# Polygenic risk, lifestyle, and cardiovascular mortality: a prospective population-based UK Biobank study

**DOI:** 10.1101/2021.02.15.21251790

**Authors:** Jae-Seung Yun, Sang-Hyuk Jung, Manu Shivakumar, Brenda Xiao, Amit V. Khera, Woong-Yang Park, Hong-Hee Won, Dokyoon Kim

## Abstract

**OBJECTIVE:** To assess the prognostic ability of polygenic risk scores (PRSs) for coronary artery disease (CAD) and type 2 diabetes mellitus (T2DM) for cardiovascular (CV) mortality, independent of traditional risk factors, and further investigate the additive effect between lifestyle behavior and PRS on CV mortality.

**DESIGN:** Prospective population-based cohort study.

**SETTING:** UK Biobank.

**PARTICIPANTS:** A total 377,909 unrelated participants of white British descent were included in the analyses from the UK Biobank cohort.

**MAIN OUTCOME MEASURES:** Genome-wide PRSs were constructed using >6 million genetic variants. We stratified patients into four PRS risk groups: low (0 to 19^th^ percentile), intermediate (20 to 79^th^ percentile), high (80 to 98^th^ percentile), and very high (99^th^ percentile). We defined a favorable and unfavorable lifestyle with four modifiable lifestyle components, including smoking, obesity, physical activity, and diet. Cox proportional hazard models were used to analyze the relationship between PRS and CV mortality with stratification by age, sex, disease status, and lifestyle behavior.

**RESULTS:** Of 377,909 UK Biobank participants having European ancestry, 3,210 (0.8%) died due to CV disease during a median follow-up of 8.9 years. CV mortality risk was significantly associated with CAD PRS (low vs. very high genetic risk groups, CAD PRS hazard ratio [HR] 2.61 [2.02 to 3.36]) and T2DM PRS (HR 2.08 [1.58 to 2.73]), respectively. These relationships remained significant even after adjustment for a comprehensive range of demographic and clinical factors. In the very high genetic risk group, adherence to an unfavorable lifestyle was further associated with a substantially increased risk of CV mortality (favorable versus unfavorable lifestyle with very high genetic risk for CAD PRS, HR 8.31 [5.12 to 13.49]; T2DM PRS, HR 5.84 [3.39 to 10.04]). Across all genetic risk groups, 32.1% of CV mortality was attributable to lifestyle behavior (population attributable fraction [PAF] 32.1% [95% CI 28.8 to 35.3%]) and 14.1% was attributable to smoking (PAF 14.1% [95% CI 12.4 to 15.7%]). There was no evidence of significant interaction between PRSs and age, sex, or lifestyle behavior in predicting the risk of CV mortality.

**CONCLUSION:** PRSs for CAD or T2DM and lifestyle behaviors are independent predictive factors for future CV mortality in the white, middle-aged population. PRS-based risk assessment could be useful to identify individuals who need intensive behavioral or therapeutic interventions to reduce the risk of CV mortality.

**Summary Box:** *What is already known on this topic:* Polygenic risk scores quantify the inherited risk conferred by the cumulative impact of common variants into a quantitative risk estimate. Previous studies primarily targeted the ability of polygenic risk scores to predict a specific disease, and only a few studies have investigated the association between genetic risk scores and cardiovascular mortality. The majority of previous analyses calculated polygenic risk scores from only a small number of genetic variants or adjusted for only a few risk factors, and no studies have examined whether the association of polygenic risk score with cardiovascular mortality differs by lifestyle behavior.

*What this study adds:* Genetic risk and lifestyle are independent predictive factors for cardiovascular mortality, even after adjustment for a comprehensive range of demographic and clinical factors. A healthy lifestyle is associated with relative risk reduction for cardiovascular mortality across all genetic risk categories, a finding that indicates the potential benefit of intensive lifestyle modification in overcoming genetic risk for cardiovascular mortality.

## Introduction

Common chronic diseases such as cardiovascular (CV) disease or diabetes represent a huge public health burden. Cardiovascular disease (CVD) is one of the leading causes of mortality worldwide; it has been estimated that CVD accounts for about 30% of all-cause mortality^1^. It is well established that type 2 diabetes mellitus (T2DM) is also a major risk factor for mortality.^2^ Moreover, people with T2DM have a higher risk of CV mortality than people without diabetes, and risk of mortality continuously increases as glycemic levels increase, even if levels do not reach those needed for a diabetes diagnosis.^3^ Thus, early screening and prevention in individuals at risk of these diseases are important strategies for reducing CV morbidity and mortality.

Traditional risk factors for common chronic diseases do not typically manifest early in life, thus it is difficult to fully identify high-risk individuals. Polygenic risk scores (PRSs) comprised of single nucleotide polymorphisms (SNPs) offer a means for early screening of and preventive interventions in common chronic diseases, including coronary artery disease (CAD) and diabetes.^4^ Many studies have demonstrated the predictive ability of PRSs to identify those with higher genetic risk of incident disease. For instance, Khera et al. suggested that PRSs measuring the cumulative genetic burden of five common diseases are well-correlated with case status.^5^ However, most previous studies have assessed PRS in terms of predicting specific diseases; the utility of PRS in predicting mortality has not yet been evaluated comprehensively.^5–7^ A few studies have shown associations between PRS and disease-specific mortality but included only a small number of SNPs. ^8–11^ Furthermore, a mortality event is a complicated outcome in which a number of potential confounding factors are involved, and the majority of previous analyses did not adjust for the comprehensive range of demographic, lifestyle, and clinical factors that can affect the risk of mortality. It is also uncertain if a healthy lifestyle can offset the genetic risk for CV mortality. Thus, it is important to evaluate the association between CV mortality and a PRS for CAD and T2DM calculated using millions of SNPs, to test this association for significance after more thorough adjustments, and to examine any differences among individuals grouped by their age, sex, disease status, or lifestyle behaviors.

The UK Biobank is a nationwide, prospective, population-based cohort of participants aged 40 to 69 and provides a variety of genetic, phenotypic, and health-related information.^12^ We primarily aimed to investigate associations between the PRSs for CAD and T2DM, lifestyle behaviors, and CV mortality risk in middle-aged UK Biobank participants of European ancestry. We further investigated the association between PRS and CV mortality according to age, sex, and disease status.

## Methods

### Study population

The UK Biobank recruited 502,505 participants aged 40 to 69 years between 2007 and 2010.^12^ At baseline, participants provided their signed consent and completed a touchscreen questionnaire, in-person interview, and physical assessment in one of 22 assessment centers across the UK. The touchscreen questionnaires covered sociodemographic factors, lifestyle behaviors, and health-related factors such as smoking status, alcohol frequency, eating habits, and medical history. Anthropometric measurements, including height, weight, waist circumference, and blood pressure, were measured by trained staff according to a standard protocol. During the baseline assessment visit, blood samples were collected and processed by standardized protocols.^13^ Only baseline assessments were used in the present study. The UK Biobank was approved by the National Research Ethics Committee (June 17, 2011 [RES reference 11/NW/0382]; extended on May 10, 2016 [RES reference 16/NW/0274]). This research using the UK Biobank Resource was approved under Application Number 33002.

### Genotyping and quality control

UK Biobank samples (version 3; March 2018) were genotyped using either the Affymetrix UK BiLEVE Axiom array or the Affymetrix UK Biobank Axiom array; these include >800,000 genotyped SNPs, 95% of which are shared between the two platforms. Imputation via IMPUTE2 was carried out centrally by UK Biobank researchers using the merged 1000 Genomes Project panel and UK 10K panel.^14^ We used genetic data from participants identified as ‘white-British’ ancestry on the basis of both self-report and principal component analysis of ancestry. We excluded individuals whose reported sex did not match with that inferred from genetic data and individuals with second-degree or closer relatives also in the Biobank. After exclusions, 377,909 individuals were eligible for the genetic analyses. After imputation, variant-level quality control (QC) was carried out by filtering SNPs on the basis of: (1) minor allele frequency <0.01 and (2) imputation quality score (INFO) <0.3. A total of 9,505,768 imputed autosomal SNPs passed the QC criteria.

### Polygenic risk scores

To generate individual genetic risk scores, we derived PRSs based on the pre-calculated weights for SNPs provided by Khera et al., which were determined using LDpred from large-scale genome-wide association study (GWAS) summary statistics.^5^ PRSs for T2DM and CAD were respectively constructed based on GWAS summary statistics from the Diabetes Genetics Replication and Meta-analysis (DIAGRAM) consortium and the Coronary Artery Disease Genome-wide Replication and Meta-analysis plus the Coronary Artery Disease Genetics (CARDIOGRAMplusC4D) consortium.^15 16^

### Ascertainment of mortality outcomes

All participants provided their consent for follow-up linkage to their death registration and health-related records. Date and underlying cause of death were obtained from death certificates provided by the National Health Service Central Register (Scotland) and the National Health Service Information Center (England and Wales). Mortality data for our UK Biobank dataset were available until November 30, 2016 for centers in Scotland and until January 31, 2018 for centers in England and Wales. Causes of death were classified using the 10^th^ revision of the International Statistical Classification of Disease (ICD-10).

### Ascertainment of variables

Information on smoking status, alcohol frequency, physical activity, eating habits, medical history, and medication use were collected via a touchscreen questionnaire or in-person interview during the baseline visit for the UK Biobank project. Height, weight, and waist circumference were assessed by trained medical staff at the same time. Regular physical activity was defined as participating in either moderate activity ≥5 days a week or vigorous activity ≥3 days a week. We used four lifestyle factors: current smoking, obesity, physical activity, and dietary pattern, as recommended by the strategic goals of the American Heart Association (AHA).^7 17 18^ Participants were categorized based on overall lifestyle scores into the following four risk subgroups: favorable (defined as having at least three healthy lifestyle factors), intermediate (having two healthy lifestyle factors), and unfavorable (having one or fewer healthy lifestyle factors). Additional definitions and details regarding lifestyle factors are provided in Supplementary Table 1.

**Table 1.** Characteristics of participants at baseline

|  | **Total**<br>**(N=377,909)** |
| --- | --- |
| **Demographics & Physical measurement** |  |
| Age (year) | 56.5 ± 7.9 |
| Sex (men) | 174,829 (46.3) |
| Systolic blood pressure (mmHg) | 140.3 ± 19.7 |
| Diastolic blood pressure (mmHg) | 82.3 ± 10.7 |
| BMI (kg/m<sup>2</sup>) | 27.4 ± 4.8 |
| Waist circumference (cm) | 90.4 ± 13.5 |
| Lifestyle behavior |  |
| Favorable | 196,450 (54.0) |
| Intermediate | 122,695 (33.8) |
| Unfavorable | 44,333 (12.2) |
| **Laboratory findings** |  |
| HbA1c (%) | 5.4 ± 0.6 |
| Estimated GFR (mL/min/1.73m<sup>2</sup>) | 78.6 ± 14.2 |
| Total cholesterol (mg/dL) | 220.7 ± 44.2 |
| Triglyceride (mg/dL) | 155.7 ± 90.6 |
| HDL cholesterol (mg/dL) | 56.1 ± 14.8 |
| LDL cholesterol (mg/dL) | 137.9 ± 33.6 |
| **Baseline major comorbidity** |  |
| Coronary artery disease | 21,678 (5.7) |
| Type 2 diabetes mellitus | 15,034 (4.2) |
| Dyslipidemia | 69,767 (18.5) |
| Hypertension | 112,350 (29.7) |
| Myocardial infarction | 9,472 (2.5) |
| Heart failure | 2,626 (0.7) |
| Ischemic stroke | 6,744 (1.8) |
| Chronic lung disease | 55,410 (14.7) |
| Chronic kidney disease | 5,766 (1.5) |
| Cancer | 45,102 (11.9) |
| Charlson comorbidity index | 0.57 ± 0.97 |

Data are n (%) or mean (SD). GFR, glomerular filtration rate

Diagnosis of prevalent CAD or T2DM at baseline was based on self-report in the in-person interview at enrollment or on diagnostic and procedure codes in electronic health records. Medical history of other major comorbidities was also collected concerning dyslipidemia, hypertension, heart failure, ischemic stroke, chronic lung disease, chronic kidney disease, and cancer. We used self-reported diagnoses, hospitalization records, and first occurrence information to define prevalent comorbidities; these definitions are presented in Supplementary Table 2. The Charlson comorbidity index (CCI), a composite comorbidity score, was calculated based on ICD-10 codes and was used to assess the severity of any underlying comorbidities.^19^ Estimated glomerular filtration rate (eGFR) was calculated using the Chronic Kidney Disease Epidemiology Collaboration (CKD-EPI) equation.^20^

### Statistical analysis

Continuous variables are reported as means with standard deviations, and categorical variables as frequencies and proportions. Baseline characteristics were compared between genetic risk groups using the chi-square test for categorical variables and ANOVA for continuous variables. The association between PRS status and mortality was investigated using Cox proportional hazards models. As in previous studies, we categorized study participants on the basis of PRS into low, intermediate, or high groups^7 18^ and further classified the top 1% of the PRS distribution as a very high risk group in light of the curve of cumulative incidence of prevalent disease over the PRS distribution (Supplementary Fig 1). Thus, participants were categorized into the following four risk subgroups: low (0 to 19^th^ percentile), intermediate (20 to 79^th^ percentile), high (80 to 98^th^ percentile), and very high (99^th^ percentile). Model 1 was adjusted for age, sex, genotyping array, and the first ten principal components (PCs) of ancestry. Model 2 was fully adjusted for the major variables associated with risk of CV mortality, such as age, sex, genotyping array, the first ten PCs, baseline blood pressure, lifestyle behavior, laboratory findings, baseline use of medications, CCI, and major comorbidities including hypertension, dyslipidemia, ischemic stroke, heart failure, cancer, chronic liver disease, chronic lung disease, and chronic kidney disease. We also calculated a P value for linear trends in regression based on the categorical risk groups. The population attributable fraction (PAF) was calculated to quantify the proportion of CV mortality in a population attributable to lifestyle behaviors. Stratified analyses were conducted using cutoffs and considered lifestyle behaviors, age at enrollment, sex, and the presence of CAD and T2DM at baseline. To investigate whether effect modification of the association between PRS and mortality occurred for lifestyle behaviors, age, sex, or disease status, we assessed multiplicative interactions between PRS and each of the stratification variables. In our survival analyses, individuals were censored according to the date of follow-up loss, the date of follow-up end (January 31, 2018 for England and Wales; November 30, 2016 for Scotland), or the date of death. Cases with missing data were excluded from modeling (Supplementary Table 3). Log minus log plots and Schoenfeld residuals were used to assess the proportional hazard assumption. All statistical tests were two-sided, and a P<0.05 was defined as statistically significant. All statistical analyses were performed using PLINK 1.9 and R (version 3.9.0).

**Figure 1.**
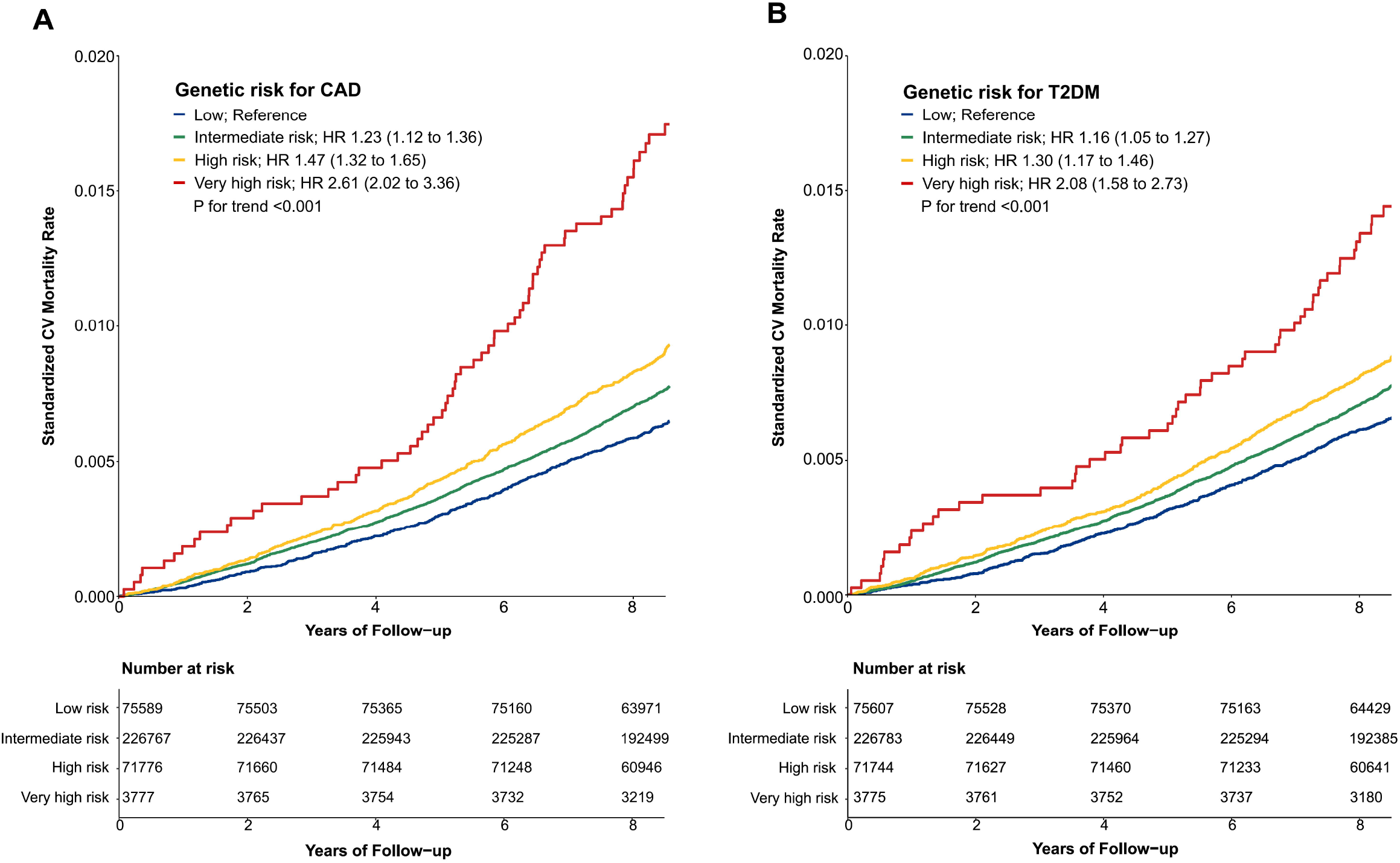
Standardized cardiovascular mortality rates according to categories of polygenic risk score for coronary artery disease (A) and type 2 diabetes mellitus (B)

### Patient and public involvement

The development of the research question or outcome measures was not informed by patients’ priorities, experience, or preferences. No patients were involved directly in the setting of the design or implementation of the study. There are no plans to involve patients in dissemination.

## Results

### Population characteristics

Baseline demographic and clinical characteristics of all 377,909 participants are presented in Table 1; characteristics for each PRS group are presented in the Supplementary Table 4. Men comprised 46.3% of participants, and the mean age at the study baseline was 56.5 years. Overall, participants at higher genetic risk for CAD or T2DM tended to have higher blood pressure, BMI, waist circumference, HbA1c, estimated glomerular filtration rate, CCI, and more frequent use of aspirin, anti-hypertensive agents, or lipid-lowering agents than those at low genetic risk.

### Association of PRS with cardiovascular mortality

During the median follow-up of 8.9 years (IQR 8.3 to 9.5 years), 15,570 participants (4.1%) died; of those deaths, 21.1% were related to CV. The overall mortality rate was 0.95 per 1000 person-years (95% CI 0.92 to 0.99). Compared with low genetic risk, higher genetic risk was associated with a higher risk of CV mortality during follow-up (P for trend <0.001) (Figure 1, Supplementary Table 5). In stepwise multivariable models, very high genetic risk for either CAD or T2DM remained significantly associated with a high risk of CV mortality, although the strength of the association was attenuated (Supplementary Table 5). In the fully-adjusted model, participants at very high genetic risk for CAD or T2DM had approximately two-fold increased risk of CV mortality relative to those with the corresponding low genetic risk. Associations between PRSs and mortality were attenuated when adjusting for PRS-specific disease status at baseline (Supplementary Table 5, model 4).

### PRS and lifestyle behaviors for cardiovascular mortality

Compared to a favorable lifestyle, an unfavorable lifestyle was associated with an increased risk of CV mortality in all genetic risk groups for CAD and T2DM alike (Figure 2, Supplementary Table 6, 7, and Supplementary Fig 2). In the groups with high and very high genetic risk for CAD, adherence to an unfavorable lifestyle was respectively associated with 4.6- and 8.3-fold increased risk of CV mortality (P value <0.001). Similarly, in the high and very high genetic risk groups for T2DM, an unfavorable lifestyle was associated with 3.9- and 5.8-fold respective increased risk of CV mortality (P value <0.001). Adherence to a favorable lifestyle was associated with reduced risk of CV mortality across all genetic risk categories (unfavorable vs. favorable lifestyle group, HR 0.33 [95% CI 0.30 to 0.36]). However, participants at very high genetic risk with a favorable lifestyle still featured high mortality risk (hazard ratio [HR] 2.82 for CAD and 1.59 for T2DM). The population attributable fraction (PAF) of favorable lifestyle behaviors for CV mortality was 32.1% (95% CI 28.8 to 35.3), and the PAF of smoking was 14.1% (95% CI 12.4 to 15.7%). There were no significant interactions between PRSs and lifestyle behaviors in predicting risk of CV mortality. Among the four lifestyle habits included in this analysis, smoking with very high genetic risk for CAD and T2DM had the strongest effect on CV mortality (Supplementary Fig 3, 4).

**Figure 2.**
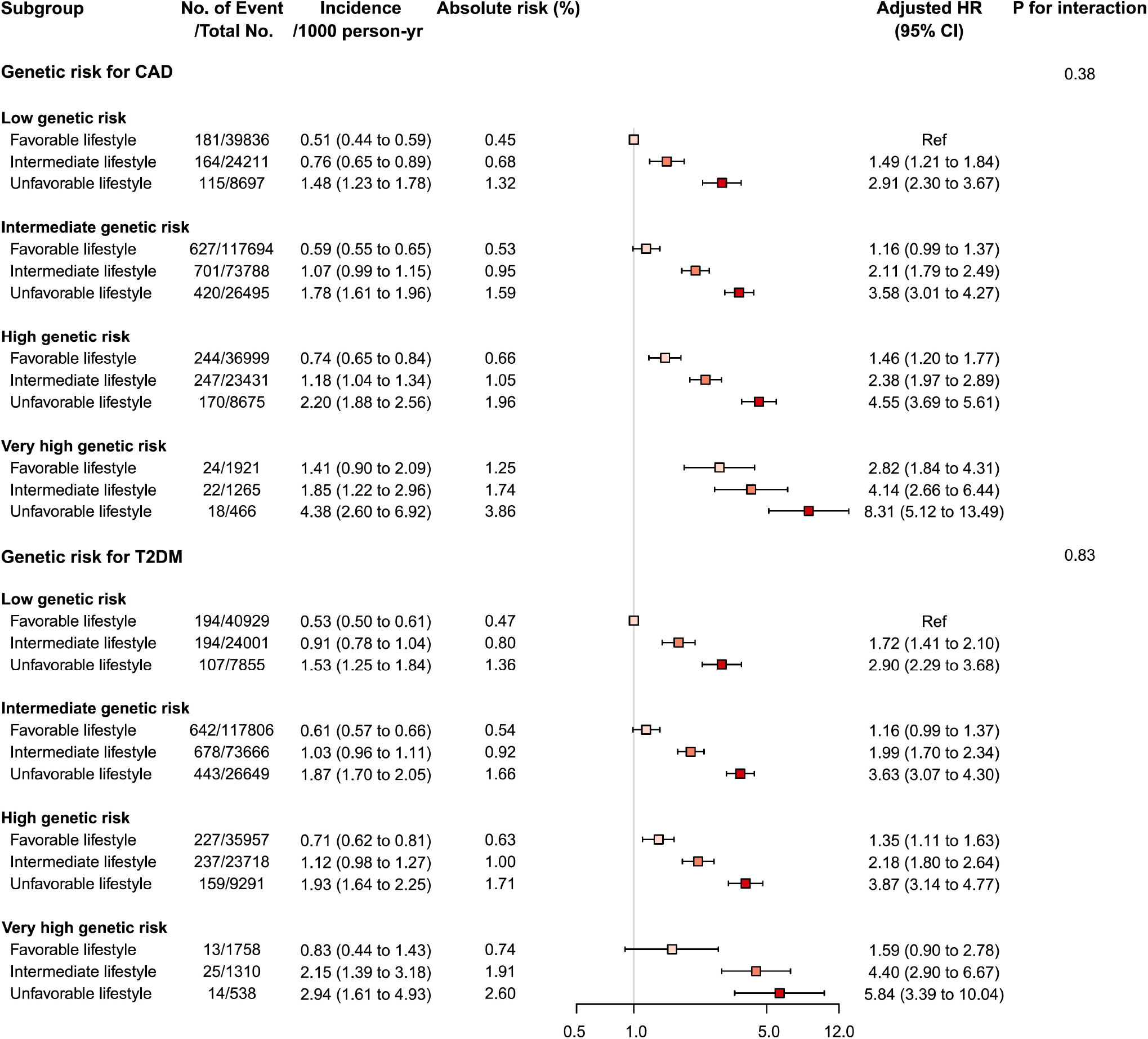
Forest plot of cardiovascular mortality according to genetic risk and lifestyle risk. Cox regression model were adjusted for age, sex, genotyping array, and first ten principal components of ancestry. P values are for testing the interaction between each genetic risk category and lifestyle category.

**Figure 3.**
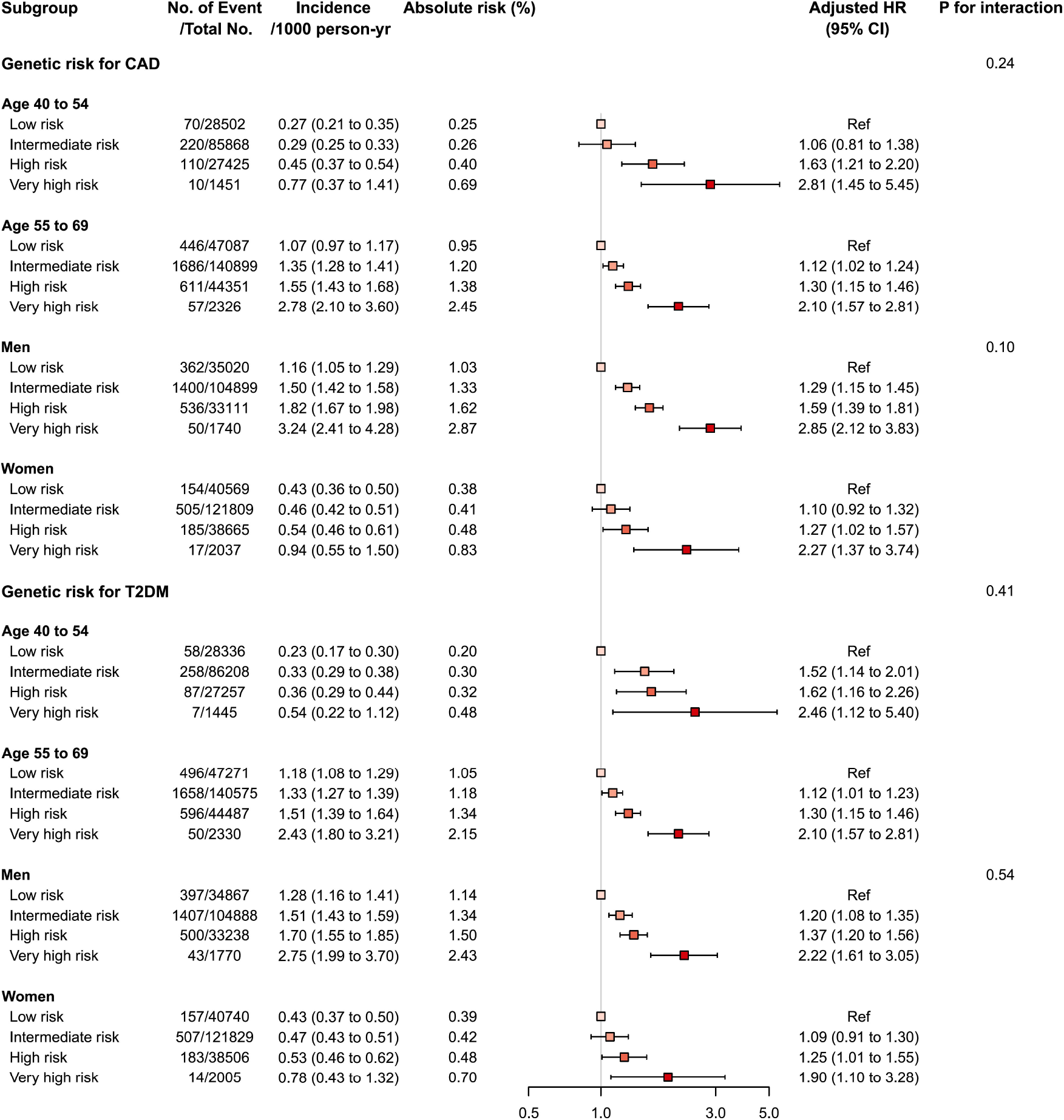
Forest plot of cardiovascular mortality according to genetic risk, age, and sex. Cox regression model were adjusted for age, sex, genotyping array, and first ten principal components of ancestry. P values are for testing the interaction between each genetic risk category, age, and sex.

**Figure 4.**
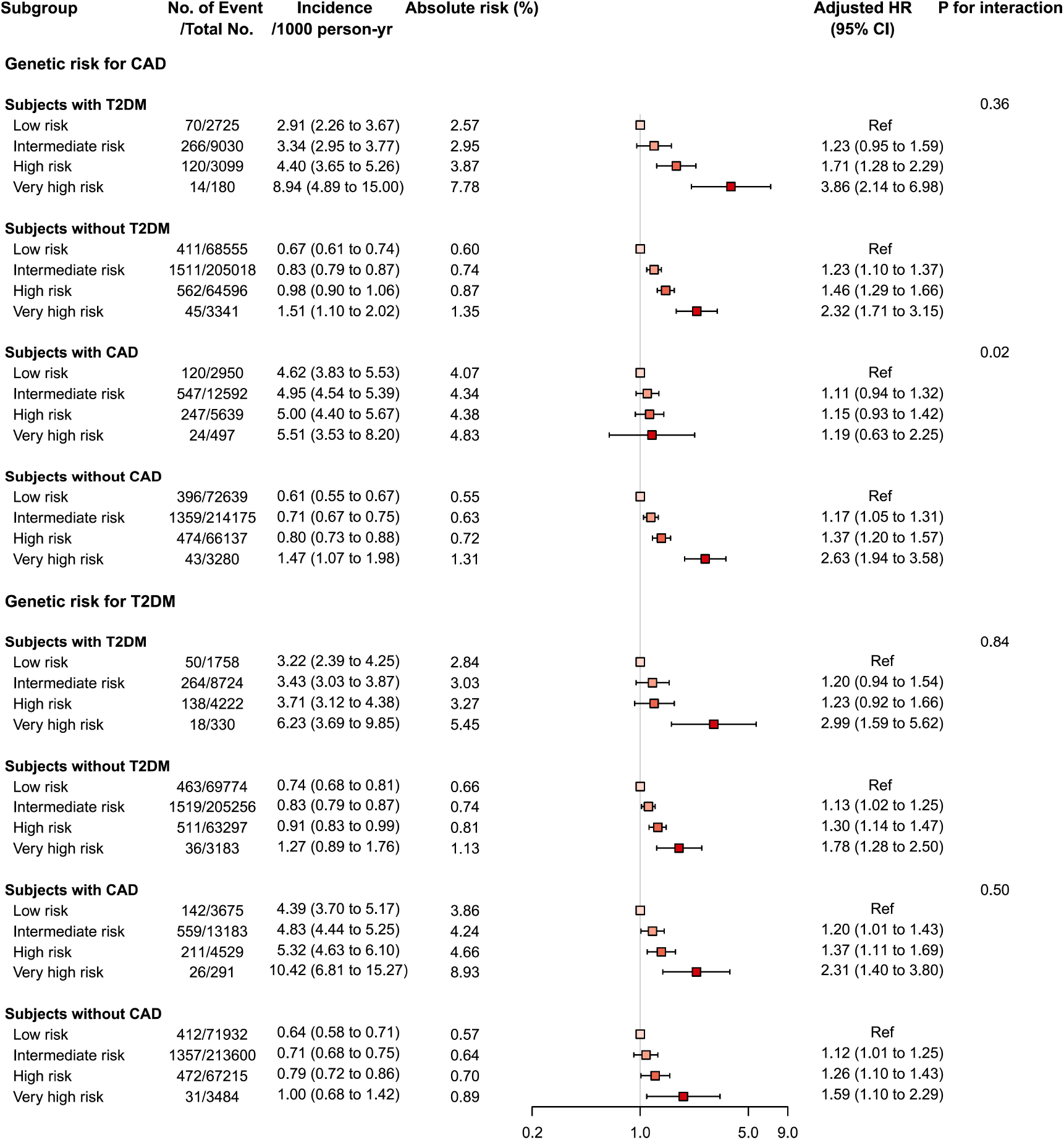
Forest plot of cardiovascular mortality according to genetic risk and PRS-related disease status. PRS, polygenic risk score Cox regression model were adjusted for age, sex, genotyping array, and first ten principal components of ancestry. P values are for testing the interaction between each genetic risk category and disease status.

### PRS, age, sex, and disease status for cardiovascular mortality

We investigated the interactions of PRS, age, and sex in the context of mortality risks and found that the association of mortality risk with PRS varied slightly by age and sex (Figure 3, Supplementary Table 8, 9, and Supplementary Fig 5). Higher genetic risks of CAD and T2DM were associated with CV mortality both in the young (<55 years) and the old (≥55 years) and were associated with increased risk of CV mortality in both sexes, with generally higher risk in men. The CAD PRS was not associated with risk of CV mortality among participants with CAD at baseline, even in those having very high genetic risk (HR 1.19 [95% CI 0.63 to 2.24], P=0.60). By contrast, a robust association between very high genetic risk for T2DM and mortality was observed among participants with CAD or T2DM at baseline (CAD PRS, HR 2.31 [1.40 to 3.80]; T2DM PRS, HR 2.99 [1.59 to 5.62], P<0.001) (Figure 4, Supplementary Table 10, and Supplementary Fig 6, 7).

## Discussion

In this prospective nationwide cohort study, we comprehensively explored the prognostic utility of PRS in predicting CV mortality. We found that the genetic risk of CAD and of T2DM were independent predictive factors for CV mortality. Furthermore, participants at very high genetic risk for CAD or T2DM, equivalent to 1% of the total population, had a 2.6- and 2.1-fold increased risk of CV mortality, respectively, relative to those at low genetic risk. These associations were attenuated but still evident even after adjustment for a wide range of mortality-related clinical and lifestyle behavior factors. In all genetic risk groups, adherence to a favorable lifestyle was associated with a reduced risk of CV mortality. In groups with very high genetic risk for CAD or T2DM, CV mortality risk remained substantially high even with a favorable lifestyle.

Previous studies have identified an association between mortality and genetic risk scores incorporating CVD-related variants. Two studies reported that CV mortality is significantly associated with a PRS calculated with a small number of variants.^10 11^ Recently, Meisner et al. described a significant association between CAD PRS and CAD-specific mortality using the UK Biobank dataset,^21^ and Damask et al. likewise reported that those with high genetic risk for CAD had 50% higher risk of major adverse cardiac events.^22^ Fewer studies have investigated the association of genetic risk of diabetes with mortality. A previous longitudinal cohort study by Leong et al. reported a borderline significant association between genetic risk for T2DM and all-cause mortality.^9^ Others have suggested that genetic risk scores associated with hyperglycemia or diabetes could not predict mortality or age of death.^21^ In the context of CVD, diabetes and hyperglycemia are well-known major risk factors for CVD and related mortality,^23^ but controversy exists regarding the effect of genetic variants associated with diabetes risk on CV mortality. Previous studies have suggested a shared genetic basis between T2DM and CAD with a complex bidirectional relationship,^24^ and several Mendelian randomization studies supported the hypothesis that genetic mechanisms linked with risk of T2DM may have causal roles in the etiology of CVD.^25 26^ Our results expand on previous research by demonstrating the significant and robust association of genetic risk of T2DM as well as CAD with mortality using a large dataset comprised of UK Biobank participants. In the model fully adjusted for established clinical CV risk factors, the HRs of PRSs were attenuated, but both genetic scores remained significant independent predictors of future CV mortality. Our finding implies that although the traditional clinical risk factors included as covariates in this study influence CV mortality, there are multiple additional pathways related to PRS that affect CV risk and mortality, such as subclinical atherosclerosis, subclinical metabolic derangement, and clinical events related to CV mortality during follow-up periods.^27 28^

In our study, we found that genetic risk and modifiable lifestyle habits were independently associated with risk of CV mortality. To the best of our knowledge, this is the first study to investigate the associations of lifestyle habits and genetic risk with CV mortality. Previous studies have mainly targeted the interaction between genetic risk and lifestyle habits for specific diseases. For example, Khera et al. suggested that both genetic factors and adherence to an unfavorable lifestyle conjointly contributed to an increased risk of CAD.^18^ Other studies reported that combined unfavorable lifestyle behaviors and genetic risk had an additive deleterious effect on the risk of developing CAD, stroke, and T2DM.^4 7 29^ The analyses in the current study showed that the risk of CV mortality was reduced by adherence to a favorable lifestyle, even in high genetic risk groups. These findings indicate the potential benefits of lifestyle modifications such as diet, weight control, regular physical activity, and smoking cessation in preventing CV mortality, regardless of genetic risk. Our analysis showed that over 30% of CV mortality might have been reduced if all participants would have adhered to favorable lifestyle behavior. In particular, patients could adjust their smoking habits, as current smoking was the most deleterious risk factor for CV mortality out of the behaviors we analyzed. The effect of lifestyle intervention in individuals at high genetic risk for CV mortality should be verified in future studies.

Several studies have reported that the effect of genetic risk on disease onset or mortality varies according to age and sex. For example, Tada et al. reported that PRS risk for CVD was higher in younger individuals than in older ones^11^, and Mars et al. suggested that higher genetic risk scores were associated with early onset of diseases including CAD and T2DM.^6^ An early onset of CVD in a high genetic risk group can be consistent with an increased risk of CV mortality in a young age group. Regarding risk and sex, while men have a higher rate of CVD incidence and mortality in the general population, the risks for CVD and its mortality have been shown to be greater in women with diabetes than in men with diabetes.^29 30^ However, we observed no significant interactions between PRS, age, and sex with regard to CV mortality. Our results indicated that the effects of PRSs on CV mortality were not stronger in women than in men, even among participants with diabetes.

This study also found that, among participants with T2DM at baseline, PRSs for T2DM and CAD were significantly associated with increased risk of CV mortality. Consistent with our findings, Cox et al. showed that a genetic risk score based on SNPs associated with CVD in patients with T2DM was associated with all-cause and CV mortality.^8^ Meanwhile, among individuals in our study with CAD at baseline, PRS for CAD was not associated with CV mortality. This finding is in line with previous results that PRSs determined using SNPs associated with CAD are not associated with recurrent CV events.^31^ Individuals with a medical history of CAD used more aggressive medications such as aspirin and lipid-lowering agents for secondary prevention; such post-event care efforts might reduce the impact of factors mediated by genetic risk for CAD on CV mortality. A previous post-hoc analysis of clinical trials has shown that high genetic risk of CAD can be mitigated by statin treatment.^32^

Several limitations of our study should be considered. First, our study could be affected by competing risk from other cause-specific mortalities, such as cancer. To account for the potential effect of competing risk of death from other causes, we adjusted for the presence at baseline of major comorbidities such as cancer, chronic lung disease, and chronic liver disease. Second, our definition of comorbidity included self-reported physician-made diagnosis, which may be incomplete and fail to include all true patients with comorbidities. However, self-reported disease status or history was obtained through a verbal interview with a trained nurse and was found to be the strongest predictor of all-cause mortality in men in a previous study.^33^ Third, lifestyle behaviors were based on a single measurement at baseline and were nonrandomized. Fourth, the predictive power of PRS can be further refined by using summary statistics from larger GWASs, which may change the estimated risk of CV mortality in individuals at high genetic risk. Finally, the association between genetic risk and CV mortality was not validated in external cohorts nor in non-European populations. Further studies with other cohorts are warranted to verify the generalizability of our findings.

In summary, our study used a large, well-phenotyped, prospective cohort drawn from the UK Biobank to assess the association between PRSs and CV mortality. We found that PRSs for CAD and T2DM are independent predictive factors for future CV mortality for this white, middle-aged cohort. We were able to adjust for various demographic, clinical, and lifestyle factors to control for potential confounding factors for mortality. The associations remained robust after adjustment for a wide range of clinical variables, including traditional CV risk factors. PRS thus has prognostic clinical utility in identifying people at risk for CVD and mortality. Furthermore, due to the considerable number of participants, we were also able to perform analyses stratified by lifestyle risk. Our findings suggest that PRS-based risk assessment could be useful among individuals with high genetic risk for CAD or T2DM who need intensive behavioral or therapeutic interventions to prevent CV mortality. PRS can stratify high risk groups associated with an increased risk of CV death as well as specific diseases at an early age or early stage of disease. This stratification could be used for early clinical support and lifelong intervention and thereby enable precision medicine. Further studies are needed to verify the efficacy of intervention or cost-effectiveness of risk assessment based on PRS.

## Data Availability

The UK Biobank dataset was obtained from the UK Biobank (Application Number 33002), and a full list of the variables are available online. Data cannot be shared publicly due to the violation of patient privacy and the absence of informed consent for data sharing.

## Acknowledgments

This work was supported by a National Research Foundation of Korea (NRF) grant funded by the Korea government (MSIT) (No. 2019R1A2C4070496) and by the National Institute of General Medical Sciences (NIGMS) R01 GM138597.

## Author Contributions

Jae-Seung Yun developed the study design, wrote the manuscript, and analyzed and interpreted data. Sang-Hyuk Jung analyzed data, interpreted data, and contributed to discussion. Manu Shivakumar analyzed data and revised the manuscript. Brenda Xiao and Woong-Yang Park revised the manuscript. Amit V. Khera, Hong-Hee Won, and Dokyoon Kim interpreted results, contributed to discussion, and revised the manuscript.

