## Supplementary materials for "Polygenic risk, lifestyle, and cardiovascular mortality: a prospective population-based UK Biobank study"

**Supplementary Table 1. Detailed definitions of lifestyle factors and lifestyle behavior**

| **Lifestyle factors** | **Component** | **Healthy lifestyle** | **Field ID of UK biobank** |
| --- | --- | --- | --- |
| Current smoking | Current smoking at baseline | Absence | 20116 |
| Obesity | BMI at baseline | <30 kg/m^2^ | 21001 |
| Physical activity | Number of days per week of physical activity 10+ minutes | Participating in moderate activity ≥5 days a week or vigorous activity ≥3 days a week | 884 (Moderate physical activity 10+ minutes) 904 (Vigorous physical activity 10+ minutes) |
| Eating habits | At least half of all following diet components was considered as a healthy lifestyle, less than half was considered as an unhealthy lifestyle | | |
|  | Fruit | ≥3 serving/day | 1309 (Fresh fruit) 1319 (Dried fruit) |
|  | Vegetable | ≥3 serving/day | 1289 (Cooked vegetables) 1299 (Salad or raw vegetables) |
|  | Whole grains | ≥3 serving/day | 1438, 1448 (Wholemeal or wholegrain bread) 1458, 1468 (Bran, oat, muesli cereal) |
|  | Fish | ≥2 serving/week | 1329 (Oily fish) 1339 (Non-oily fish) |
|  | Dairy | ≥2.5 serving/week | 1408 (Cheese) 1418 (Milk) |
|  | Refined grains | ≤1.5 serving/week | 1438, 1448 (Wholemeal or wholegrain bread) 1458, 1468 (Bran, oat, muesli cereal) |
|  | Processed meats | ≤1 serving/week | 1349 (Processed meat) 3680 (Age when last ate any kind of meat, 0 if indicated having never eaten meat) |
|  | Unprocessed meats | ≤1.5 serving/week | 1359 (Poultry) 1369 (Beef) 1379 (Lamb) 1389 (Pork) 3680 (Age when last ate any kind of meat, 0 if indicated having never eaten meat) |
|  | Sugar-sweetened beverages | ≤1 serving/week | 6144 (Never eats sugar or foods/drinks containing sugar) |
| Lifestyle behavior | Favorable | Having at least three healthy lifestyle factors | |
|  | Intermediate | Having two healthy lifestyle factors | |
|  | Unfavorable | Having one or fewer healthy lifestyle factor | |

**Supplementary Table 2. Detailed definitions of comorbidities**

| **Disease** | **Path** | **Field ID** | **Code** |
| --- | --- | --- | --- |
| Dyslipidemia | Verbal interview | 20002 | 1473 |
|  | First occurrence before enrollment | 130815, 130816 | E78 |
|  | Medication | 6177 | Cholesterol lowering medication |
| Hypertension | Verbal interview | 20002 | 1065, 1072 |
|  | Touchscreen | 6150 | High blood pressure |
|  | First occurrence before enrollment | 131286, 131287, 131288, 131289, 131290, 131291, 131292, 131293, 131294, 131295 | I10, I11, I12, I13, I15 |
|  | Medication | 6177 | Blood pressure medication |
| Type 2 diabetes mellitus | Verbal interview | 20002 | 1220, 1223 |
|  | Touchscreen | 2443 | Yes |
|  | First occurrence before enrollment | 130708, 130709, 130714, 130715 | E11, E14 |
|  | Medication | 20003 | Insulin: 1140883066  Metformin: 1140884600, 1141189090  Sulfonylurea: 1141152590, 1140874744, 1140874718, 1141156984  Acarbose: 1140868902  Thiazolidinedione: 1141171646  Meglitinide: 1141168660, 1141173882 |
|  | HbA1c at baseline | 30750 |  |
|  | Verbal interview for exclusion type 1 diabetes | 20002 | 1222 |
|  | First occurrence for exclusion type 1 diabetes | 130706, 130707 | E10 |
| Coronary artery disease | Verbal interview | 20002 | 1074, 1075 |
|  | Touchscreen | 6150 | Heart attack, angina |
|  | First occurrence before enrollment | 131296, 131297, 131298, 131299, 131300, 131301, 131302, 131303, 131304, 131305, 131306, 131307 | I20, I21, I22, I23, I24, I25 |
| Heart failure | Verbal interview | 20002 | 1076, 1079 |
|  | First occurrence before enrollment | 131354, 131355 | I50 |
|  | Hospital inpatient | 41270 | I11, I110, I113, I132, I255, I420, I421, I422, I425, I428, I50, I500, I501, IO509 |
|  |  | 41271 | 4254, 4280, 4281, 4289 |
| Ischemic stroke | Verbal interview | 20002 | 1081, 1082, 1583 |
|  | Touchscreen | 6150 | Stroke |
|  | First occurrence before enrollment | 131366, 131367, 131368, 131369 | I63, I64 |
| Chronic lung disease | Verbal interview | 20002 | 1111, 1112, 1113, 1114, 1115 |
|  | First occurrence before enrollment | 131484, 131485, 131486, 131487, 131488, 131489, 131490, 131491, 131492, 131493, 131494, 131495, 131496, 131497, 131498, 131499 | J40-J47, J60-J64, J66, J67 |
| Chronic liver disease | Verbal interview | 20002 | 1155, 1156, 1157, 1158 |
|  | First occurrence before enrollment | 130200, 130201, 131406, 131407, 131658, 131659, 131660, 131661, 131662, 131663, 131664, 131665, 131670, 131671 | B18, I85, K70-73, K76 |
| Chronic kidney disease | Verbal interview | 20002 | 1192, 1193, 1194 |
|  | First occurrence before enrollment | 132030, 132031, 132032, 132033 | N18, N19 |
| Cancer | Verbal interview | 20001 | 1071, 1085 |
|  | First occurrence before enrollment | 40006 | C00-C97 |

**Supplementary Table 3. Number of cases with missing data for each variable**

| **Variable** | **n (%)** |
| --- | --- |
| Total | 377,909 |
| Systolic blood pressure | 341 (0.1) |
| Diastolic blood pressure | 339 (0.1) |
| Height | 807 (0.2) |
| Weight | 1,058 (0.3) |
| Body mass index | 1,192 (0.3) |
| Waist circumference | 626 (0.2) |
| Current smoking | 1331 (0.4) |
| Alcohol frequency | 336 (0.1) |
| Physical activity | 1,686 (0.4) |
| Lifestyle behavior | 14,431 (3.8) |
| Total cholesterol | 17,588 (4.7) |
| Triglyceride | 17,880 (4.7) |
| HDL-cholesterol | 48,177 (12.7) |
| LDL-cholesterol | 18,257 (4.8) |
| Creatinine | 17,772 (4.7) |
| C-reactive protein | 18,382 (4.9) |
| AST | 18,914 (5.0) |
| ALT | 17,721 (4.7) |
| Glucose | 48,393 (12.8) |
| HbA1c | 17,768 (4.7) |
| Lipoprotein(a) | 91,389 (24.2) |
| White blood cell | 11,203 (3.0) |
| Hemoglobin | 11,199 (3.0) |
| Platelet | 11,202 (3.0) |
| Albumin | 48,029 (12.7) |
| Total bilirubin | 19,094 (5.1) |
| AST, Aspartate aminotransferase; ALT, Alanine aminotransferase | |

**Supplementary Table 4. Characteristics of participants at baseline according to categories of PRS for CAD and T2DM**

|  | **Total** | **PRS for CAD** | | | | | **PRS for T2DM** | | | | |
| --- | --- | --- | --- | --- | --- | --- | --- | --- | --- | --- | --- |
|  |  | **Low** | **Intermediate** | **High** | **Very high** | **P value** | **Low** | **Intermediate** | **High** | **Very high** | **P value** |
|  |  | **0st-19th percentile** | **20st-79th percentile** | **80st-98th percentile** | **99th percentile** |  | **0st-19th percentile** | **20st-79th percentile** | **80st-98th percentile** | **99th percentile** |  |
|  | **(N=377,909)** | **(N=75,589)** | **(N=226,767)** | **(N=71,776)** | **(N=3,777)** |  | **(N=75,607)** | **(N=226,783)** | **(N=71,744)** | **(N=3,775)** |  |
| **Demographics & Physical measurement** |  |  |  |  |  |  |  |  |  |  |  |
| Age (years) | 56.5 ± 7.9 | 56.5 ± 7.9 | 56.5 ± 8.0 | 56.4 ± 7.9 | 56.3 ± 7.9 | 0.002 | 56.5 ± 7.9 | 56.5 ± 8.0 | 56.5 ± 7.9 | 56.3 ± 7.9 | 0.05 |
| Men | 174,829 (46.3) | 35,020 (46.3) | 104,958 (46.3) | 33,111 (46.1) | 1,740 (46.1) | 0.86 | 34,867 (46.1) | 104,954 (46.3) | 33,238 (46.3) | 1,770 (46.9) | 0.71 |
| Systolic blood pressure (mmHg) | 140.3 ± 19.7 | 139.7 ± 19.5 | 140.3 ± 19.7 | 140.7 ± 19.7 | 141.2 ± 19.7 | <0.001 | 139.5 ± 19.7 | 140.3 ± 19.6 | 141.1 ± 19.6 | 142.3 ± 19.4 | <0.001 |
| Diastolic blood pressure (mmHg) | 82.3 ± 10.7 | 82.2 ± 10.6 | 82.3 ± 10.7 | 82.4 ± 10.7 | 82.6 ± 10.8 | 0.005 | 81.9 ± 10.6 | 82.3 ± 10.7 | 82.7 ± 10.6 | 83.5 ± 10.6 | <0.001 |
| BMI (kg/m^2^) | 27.4 ± 4.8 | 27.3 ± 4.7 | 27.4 ± 4.7 | 27.5 ± 4.8 | 27.7 ± 4.9 | <0.001 | 27.0 ± 4.6 | 27.4 ± 4.8 | 27.8 ± 4.9 | 28.5 ± 5.1 | <0.001 |
| Waist circumference (cm) | 90.4 ± 13.5 | 90.2 ± 13.5 | 90.4 ± 13.5 | 90.5 ± 13.6 | 90.9 ± 13.7 | <0.001 | 89.3 ± 13.3 | 90.4 ± 13.5 | 91.5 ± 13.6 | 93.1 ± 13.7 | <0.001 |
| Lifestyle behavior |  |  |  |  |  | <0.001 |  |  |  |  | <0.001 |
| Favorable | 196,450 (54.0) | 39,836 (54.8) | 117,694 (54.0) | 36,999 (53.5) | 1,921 (52.6) |  | 40,929 (56.2) | 117,806 (54.0) | 35,957 (52.1) | 1,758 (48.8) |  |
| Intermediate | 122,695 (33.8) | 24,211 (33.3) | 73,788 (33.9) | 23,431 (33.9) | 1,265 (34.6) |  | 24,001 (33.0) | 73,666 (33.8) | 23,718 (34.4) | 1,310 (36.3) |  |
| Unfavorable | 44,333 (12.2) | 8,697 (12.0) | 26,495 (12.2) | 8,675 (12.6) | 466 (12.8) |  | 7,855 (10.8) | 26,649 (12.2) | 9,291 (13.5) | 538 (14.9) |  |
| **Laboratory findings** |  |  |  |  |  |  |  |  |  |  |  |
| HbA1c (%) | 5.4 ± 0.6 | 5.4 ± 0.6 | 5.4 ± 0.6 | 5.5 ± 0.6 | 5.5 ± 0.6 | <0.001 | 5.4 ± 0.5 | 5.4 ± 0.6 | 5.5 ± 0.7 | 5.6 ± 0.8 | <0.001 |
| Estimated GFR (ml/min/1.73m^2^) | 78.6 ± 14.2 | 78.6 ± 14.1 | 78.5 ± 14.3 | 78.6 ± 14.3 | 78.4 ± 14.4 | 0.95 | 78.5 ± 14.0 | 78.5 ± 14.2 | 78.7 ± 14.4 | 79.0 ± 14.9 | <0.001 |
| Total cholesterol (mmol/L) | 5.72 ± 1.15 | 5.69 ± 1.12 | 5.72 ± 1.15 | 5.74 ± 1.17 | 5.76 ± 1.23 | <0.001 | 5.73 ± 1.12 | 5.72 ± 1.15 | 5.70 ± 1.17 | 5.69 ± 1.22 | <0.001 |
| Triglyceride (mmol/L) | 1.76 ± 1.02 | 1.75 ± 1.01 | 1.76 ± 1.02 | 1.76 ± 1.03 | 1.76 ± 1.08 | 0.08 | 1.68 ± 0.97 | 1.76 ± 1.02 | 1.84 ± 1.07 | 1.94 ± 1.14 | <0.001 |
| HDL-cholesterol (mmol/L) | 1.45 ± 0.38 | 1.46 ± 0.38 | 1.45 ± 0.38 | 1.44 ± 0.38 | 1.43 ± 0.38 | <0.001 | 1.48 ± 0.38 | 1.45 ± 0.38 | 1.42 ± 0.38 | 1.39 ± 0.38 | <0.001 |
| LDL-cholesterol (mmol/L) | 3.57 ± 0.87 | 3.54 ± 0.85 | 3.57 ± 0.87 | 3.60 ± 0.89 | 3.63 ± 0.93 | <0.001 | 3.57 ± 0.85 | 3.57 ± 0.87 | 3.57 ± 0.89 | 3.57 ± 0.92 | 0.11 |
| **Baseline major comorbidity** |  |  |  |  |  |  |  |  |  |  |  |
| Coronary artery disease | 21,678 (5.7) | 2,950 (3.9) | 12,592 (5.6) | 5,639 (7.9) | 497 (13.2) | <0.001 | 3,675 (4.9) | 13,183 (5.8) | 4,529 (6.3) | 291 (7.7) | <0.001 |
| Type 2 diabetes mellitus | 15,034 (4.2) | 2,725 (3.8) | 9,030 (4.2) | 3,099 (4.6) | 180 (5.1) | <0.001 | 1,758 (2.5) | 8,724 (4.1) | 4,222 (6.3) | 330 (9.4) | <0.001 |
| Dyslipidemia | 69,767 (18.5) | 11,771 (15.6) | 41,575 (18.3) | 15,300 (21.3) | 1,121 (29.7) | <0.001 | 12,222 (16.2) | 41,816 (18.4) | 14,847 (20.7) | 882 (23.4) | <0.001 |
| Hypertension | 112,350 (29.7) | 20,962 (27.7) | 67,299 (29.7) | 22,733 (31.7) | 1,356 (35.9) | <0.001 | 20,087 (26.6) | 67,451 (29.7) | 23,404 (32.6) | 1,408 (37.3) | <0.001 |
| Myocardial infarction | 9,472 (2.5) | 1,116 (1.5) | 5,391 (2.4) | 2,705 (3.8) | 260 (6.9) | <0.001 | 1,510 (2.0) | 5,822 (2.6) | 2,007 (2.8) | 133 (3.5) | <0.001 |
| Heart failure | 2,626 (0.7) | 410 (0.5) | 1,530 (0.7) | 632 (0.9) | 54 (1.4) | <0.001 | 442 (0.6) | 1,607 (0.7) | 547 (0.8) | 30 (0.8) | <0.001 |
| Ischemic stroke | 6,744 (1.8) | 1,206 (1.6) | 4,107 (1.8) | 1,355 (1.9) | 76 (2.0) | <0.001 | 1,225 (1.6) | 4,030 (1.8) | 1,399 (1.9) | 90 (2.4) | <0.001 |
| Chronic lung disease | 55,410 (14.7) | 11,150 (14.8) | 33,251 (14.7) | 10,471 (14.6) | 538 (14.2) | 0.73 | 10,422 (13.8) | 33,467 (14.8) | 10,931 (15.2) | 590 (15.6) | <0.001 |
| Chronic kidney disease | 5,766 (1.5) | 1,050 (1.4) | 3,506 (1.5) | 1,138 (1.6) | 72 (1.9) | 0.002 | 1,023 (1.4) | 3,492 (1.5) | 1,174 (1.6) | 77 (2.0) | <0.001 |
| Cancer | 45,102 (11.9) | 9,318 (12.3) | 26,988 (11.9) | 8,393 (11.7) | 403 (10.7) | <0.001 | 9,079 (12.0) | 27,007 (11.9) | 8,572 (11.9) | 444 (11.8) | 0.88 |
| Charlson comorbidity index | 0.57 ± 0.97 | 0.55 ± 0.95 | 0.57 ± 0.97 | 0.59 ± 0.99 | 0.60 ± 0.99 | <0.001 | 0.53 ± 0.94 | 0.57 ± 0.97 | 0.61 ± 1.00 | 0.66 ± 1.04 | <0.001 |

Data are n (%) or mean (SD). PRS, polygenic risk score; CAD, coronary artery disease; T2DM, type 2 diabetes mellitus; GFR, glomerular filtration rate

**Supplementary Table 5. Hazard ratios and 95% confidential intervals for the cardiovascular mortality using Cox proportional regression model**

|  |  | **CV morality** | | | | | | | | | | | | | | | | |
| --- | --- | --- | --- | --- | --- | --- | --- | --- | --- | --- | --- | --- | --- | --- | --- | --- | --- | --- |
|  |  | **No. of Events/  Total No.** | **Incidence rate per 1000 person-year (95% CI)** | **Absolute risk (%)** | **Crude** | | **Model 1** | | **Model 2** | | **Model 3** | | **Model 4** | | **Model 5** | | **Model 6** | |
|  |  |  |  |  | **HR (95% CI)** | **P value** | **HR (95% CI)** | **P value** | **HR (95% CI)** | **P value** | **HR (95% CI)** | **P value** | **HR (95% CI)** | **P value** | **HR (95% CI)** | **P value** | **HR (95% CI)** | **P value** |
| **CAD PRS** | Low risk | 516/75589 | 0.77 (0.70 to 0.84) | 0.68 | Ref |  | Ref |  | Ref |  | Ref |  | Ref |  | Ref |  | Ref |  |
|  | Intermediate risk | 1906/226767 | 0.94 (0.90 to 0.99) | 0.84 | 1.23 (1.12 to 1.36) | <0.001 | 1.23 (1.12 to 1.36) | <0.001 | 1.19 (1.05 to 1.34) | 0.004 | 1.24 (1.06 to 1.44) | 0.005 | 1.17 (1.01 to 1.36) | 0.04 | 1.17 (1.01 to 1.36) | 0.04 | 1.16 (0.99 to 1.35) | 0.05 |
|  | High risk | 721/71776 | 1.12 (1.05 to 1.21) | 1.00 | 1.47 (1.32 to 1.65) | <0.001 | 1.50 (1.33 to 1.68) | <0.001 | 1.49 (1.30 to 1.71) | <0.001 | 1.54 (1.29 to 1.83) | <0.001 | 1.38 (1.16 to 1.65) | <0.001 | 1.39 (1.16 to 1.65) | <0.001 | 1.37 (1.15 to 1.63) | <0.001 |
|  | Very high risk | 67/3777 | 2.00 (1.55 to 2.54) | 1.77 | 2.61 (2.02 to 3.36) | <0.001 | 2.68 (2.08 to 3.46) | <0.001 | 2.84 (2.10 to 3.84) | <0.001 | 2.24 (1.40 to 3.59) | <0.001 | 1.80 (1.12 to 2.90) | 0.01 | 1.96 (1.22 to 3.14) | 0.005 | 1.91 (1.19 to 3.07) | 0.007 |
| **T2DM PRS** | Low risk | 554/75607 | 0.82 (0.75 to 0.89) | 0.73 | Ref |  | Ref |  | Ref |  | Ref |  | Ref |  | Ref |  | Ref |  |
|  | Intermediate risk | 1916/226783 | 0.95 (0.91 to 0.99) | 0.84 | 1.16 (1.05 to 1.27) | 0.003 | 1.17 (1.07 to 1.29) | <0.001 | 1.16 (1.03 to 1.30) | 0.01 | 1.24 (1.07 to 1.44) | 0.004 | 1.20 (1.03 to 1.39) | 0.02 | 1.17 (1.01 to 1.36) | 0.04 | 1.17 (1.01 to 1.36) | 0.04 |
|  | High risk | 683/71744 | 1.07 (0.99 to 1.15) | 0.95 | 1.30 (1.17 to 1.46) | <0.001 | 1.33 (1.19 to 1.49) | <0.001 | 1.26 (1.10 to 1.45) | <0.001 | 1.39 (1.16 to 1.65) | <0.001 | 1.32 (1.11 to 1.58) | 0.002 | 1.26 (1.06 to 1.50) | 0.01 | 1.26 (1.06 to 1.50) | 0.01 |
|  | Very high risk | 57/3775 | 1.70 (1.29 to 2.21) | 1.51 | 2.08 (1.58 to 2.73) | <0.001 | 2.16 (1.64 to 2.83) | <0.001 | 2.09 (1.51 to 2.90) | <0.001 | 2.16 (1.43 to 3.27) | <0.001 | 2.03 (1.34 to 3.07) | <0.001 | 1.83 (1.21 to 2.78) | 0.004 | 1.82 (1.20 to 2.76) | 0.005 |

Model 1: Age + sex + genotyping array + first ten PC

Model 2: Model 1 + systolic BP + diastolic BP + BMI + waist circumference + current smoking + alcohol frequency + physical activity + eating habit

Model 3: Model 2 + HbA1c + total cholesterol + triglyceride + HDL cholesterol + LDL cholesterol + lipoprotein (a) + estimated GFR + AST + ALT + Total bilirubin + albumin + white blood cell + hemoglobin + platelet + C-reactive protein

Model 4: Model 3 + PRS specific disease

Model 5: Model 3 + hypertension + dyslipidemia + heart failure + cancer + chronic kidney disease + stroke + chronic lung disease + chronic liver disease + Charlson comorbidity index

Model 6: Model 4 + aspirin + lipid-lowering agent + anti-hypertensive agent

CAD, coronary artery disease; PRS polygenic risk score; T2DM, type 2 diabetes; PC, principal component; BP, blood pressure; GFR, glomerular filtration rate; AST, Aspartate aminotransferase; ALT, Alanine aminotransferase

CAD PRS and T2DM PRS of Model 4 was adjusted for CAD and T2DM, respectively.

**Supplementary Table 6. Hazard ratios and 95% confidential intervals for the cardiovascular mortality according to genetic risk for CAD and lifestyle behaviors**

|  |  | **No. of Events/  Total No.** | **Incidence/ 1000 person-year** | **Absolute risk (%)** | **CV mortality** | | | | | |
| --- | --- | --- | --- | --- | --- | --- | --- | --- | --- | --- |
|  |  |  |  |  | **Crude** | | **Model 1** | | **Model 2** | |
|  |  |  |  |  | **HR (95% CI)** | **P value** | **HR (95% CI)** | **P value** | **HR (95% CI)** | **P value** |
| **Low CAD genetic risk** | **Favorable lifestyle** | 181/39836 | 0.51 (0.44 to 0.59) | 0.45 | Ref |  | Ref |  | Ref |  |
|  | **Intermediate lifestyle** | 164/24211 | 0.76 (0.65 to 0.89) | 0.68 | 1.49 (1.20 to 1.84) | <0.001 | 1.49 (1.21 to 1.84) | <0.001 | 1.44 (1.10 to 1.88) | <0.001 |
|  | **Unfavorable lifestyle** | 115/8697 | 1.48 (1.23 to 1.78) | 1.32 | 2.90 (2.30 to 3.67) | <0.001 | 2.91 (2.30 to 3.67) | <0.001 | 2.04 (1.50 to 2.77) | <0.001 |
| **Intermediate CAD genetic risk** | **Favorable lifestyle** | 627/117694 | 0.59 (0.55 to 0.65) | 0.53 | 1.17 (0.99 to 1.38) | 0.06 | 1.16 (0.99 to 1.37) | 0.07 | 1.23 (0.99 to 1.53) | 0.05 |
|  | **Intermediate lifestyle** | 701/73788 | 1.07 (0.99 to 1.15) | 0.95 | 2.09 (1.77 to 2.46) | <0.001 | 2.11 (1.79 to 2.49) | <0.001 | 1.79 (1.45 to 2.22) | <0.001 |
|  | **Unfavorable lifestyle** | 420/26495 | 1.78 (1.61 to 1.96) | 1.59 | 3.49 (2.93 to 4.15) | <0.001 | 3.58 (3.01 to 4.27) | <0.001 | 2.43 (1.93 to 3.06) | <0.001 |
| **High CAD genetic risk** | **Favorable lifestyle** | 244/36999 | 0.74 (0.65 to 0.84) | 0.66 | 1.45 (1.20 to 1.76) | <0.001 | 1.46 (1.20 to 1.77) | <0.001 | 1.52 (1.19 to 1.95) | <0.001 |
|  | **Intermediate lifestyle** | 247/23431 | 1.18 (1.04 to 1.34) | 1.05 | 2.32 (1.91 to 2.81) | <0.001 | 2.38 (1.97 to 2.89) | <0.001 | 1.91 (1.49 to 2.45) | <0.001 |
|  | **Unfavorable lifestyle** | 170/8675 | 2.20 (1.88 to 2.26) | 1.96 | 4.30 (3.49 to 5.30) | <0.001 | 4.55 (3.69 to 5.61) | <0.001 | 2.77 (2.10 to 3.67) | <0.001 |
| **Very high CAD genetic risk** | **Favorable lifestyle** | 24/1921 | 1.41 (0.90 to 2.09) | 1.25 | 2.75 (1.80 to 4.22) | <0.001 | 2.82 (1.84 to 4.31) | <0.001 | 2.27 (1.18 to 4.35) | 0.01 |
|  | **Intermediate lifestyle** | 22/1265 | 1.85 (1.22 to 2.96) | 1.74 | 3.82 (2.46 to 5.95) | <0.001 | 4.14 (2.66 to 6.44) | <0.001 | 2.13 (1.11 to 4.09) | 0.02 |
|  | **Unfavorable lifestyle** | 18/466 | 4.38 (2.60 to 6.92) | 3.86 | 8.59 (5.29 to 13.94) | <0.001 | 8.31 (5.12 to 13.49) | <0.001 | 5.55 (2.80 to 11.01) | <0.001 |

Model 1: Age + sex + genotyping array + first ten PC

Model 2: Age + sex + genotyping array + first ten PC + systolic BP + diastolic BP + BMI + waist circumference + current smoking + alcohol frequency + physical activity + eating habit

+ HbA1c + total cholesterol + triglyceride + HDL cholesterol + LDL cholesterol + lipoprotein (a) + estimated GFR + AST + ALT + Total bilirubin + albumin + white blood cell + hemoglobin + platelet + C-reactive protein

+ PRS specific disease + hypertension + dyslipidemia + heart failure + cancer + chronic kidney disease + stroke + chronic lung disease + chronic liver disease + Charlson comorbidity index

+ aspirin + lipid-lowering agent + anti-hypertensive agent

CAD, coronary artery disease; PRS polygenic risk score; PC, principal component; BP, blood pressure; GFR, glomerular filtration rate; AST, Aspartate aminotransferase; ALT, Alanine aminotransferase

**Supplementary Table 7. Hazard ratios and 95% confidential intervals for the cardiovascular mortality according to genetic risk for T2DM and lifestyle behaviors**

|  |  | **No. of Events/  Total No.** | **Incidence/ 1000 person-year** | **Absolute risk (%)** | **CV mortality** | | | | | |
| --- | --- | --- | --- | --- | --- | --- | --- | --- | --- | --- |
|  |  |  |  |  | **Crude** | | **Model 1** | | **Model 2** | |
|  |  |  |  |  | **HR (95% CI)** | **P value** | **HR (95% CI)** | **P value** | **HR (95% CI)** | **P value** |
| **Low T2DM genetic risk** | **Favorable lifestyle** | 194/40929 | 0.53 (0.50 to 0.61) | 0.47 | Ref |  | Ref |  | Ref |  |
|  | **Intermediate lifestyle** | 194/24001 | 0.91 (0.78 to 1.04) | 0.80 | 1.70 (1.40 to 2.08) | <0.001 | 1.72 (1.41 to 2.10) | <0.001 | 1.43 (1.10 to 1.84) | 0.006 |
|  | **Unfavorable lifestyle** | 107/7855 | 1.53 (1.25 to 1.84) | 1.36 | 2.86 (2.26 to 3.62) | <0.001 | 2.90 (2.29 to 3.68) | <0.001 | 1.77 (1.29 to 2.42) | <0.001 |
| **Intermediate T2DM genetic risk** | **Favorable lifestyle** | 642/117806 | 0.61 (0.57 to 0.66) | 0.54 | 1.15 (0.98 to 1.35) | 0.09 | 1.16 (0.99 to 1.37) | 0.06 | 1.19 (0.97 to 1.46) | 0.09 |
|  | **Intermediate lifestyle** | 678/73666 | 1.03 (0.96 to 1.11) | 0.92 | 1.94 (1.66 to 2.28) | <0.001 | 1.99 (1.70 to 2.34) | <0.001 | 1.64 (1.34 to 2.02) | <0.001 |
|  | **Unfavorable lifestyle** | 443/26649 | 1.87 (1.70 to 2.05) | 1.66 | 3.51 (2.96 to 4.15) | <0.001 | 3.63 (3.07 to 4.30) | <0.001 | 2.35 (1.89 to 2.93) | <0.001 |
| **High T2DM genetic risk** | **Favorable lifestyle** | 227/35957 | 0.71 (0.62 to 0.81) | 0.63 | 1.34 (1.10 to 1.62) | 0.003 | 1.35 (1.11 to 1.63) | 0.002 | 1.28 (1.01 to 1.63) | 0.04 |
|  | **Intermediate lifestyle** | 237/23718 | 1.12 (0.98 to 1.27) | 1.00 | 2.11 (1.75 to 2.55) | <0.001 | 2.18 (1.80 to 2.64) | <0.001 | 1.76 (1.39 to 2.24) | <0.001 |
|  | **Unfavorable lifestyle** | 159/9291 | 1.93 (1.64 to 2.25) | 1.71 | 3.62 (2.94 to 4.47) | <0.001 | 3.87 (3.14 to 4.77) | <0.001 | 2.43 (1.85 to 3.20) | <0.001 |
| **Very high T2DM genetic risk** | **Favorable lifestyle** | 13/1758 | 0.83 (0.44 to 1.43) | 0.74 | 1.57 (0.90 to 2.76) | 0.11 | 1.59 (0.90 to 2.78) | 0.11 | 1.16 (0.54 to 2.50) | 0.70 |
|  | **Intermediate lifestyle** | 25/1310 | 2.15 (1.39 to 3.18) | 1.91 | 4.05 (2.67 to 6.14) | <0.001 | 4.40 (2.90 to 6.67) | <0.001 | 2.49 (1.40 to 4.42) | 0.03 |
|  | **Unfavorable lifestyle** | 14/538 | 2.94 (1.61 to 4.93) | 2.60 | 5.53 (3.21 to 9.50) | <0.001 | 5.84 (3.39 to 10.04) | <0.001 | 4.34 (2.27 to 8.30) | <0.001 |

Model 1: Age + sex + genotyping array + first ten PC

Model 2: Age + sex + genotyping array + first ten PC + systolic BP + diastolic BP + BMI + waist circumference + current smoking + alcohol frequency + physical activity + eating habit

+ HbA1c + total cholesterol + triglyceride + HDL cholesterol + LDL cholesterol + lipoprotein (a) + estimated GFR + AST + ALT + Total bilirubin + albumin + white blood cell + hemoglobin + platelet + C-reactive protein

+ PRS specific disease + hypertension + dyslipidemia + heart failure + cancer + chronic kidney disease + stroke + chronic lung disease + chronic liver disease + Charlson comorbidity index

+ aspirin + lipid-lowering agent + anti-hypertensive agent

T2DM, type 2 diabetes mellitus; PRS polygenic risk score; PC, principal component; BP, blood pressure; GFR, glomerular filtration rate; AST, Aspartate aminotransferase; ALT, Alanine aminotransferase

**Supplementary Table 8. Hazard ratios and 95% confidential intervals for the cardiovascular mortality according to genetic risk and age category**

| **CV mortality** | | **Age 40-54** | | | | | | | | | **Age 55-69** | | | | | | | | |
| --- | --- | --- | --- | --- | --- | --- | --- | --- | --- | --- | --- | --- | --- | --- | --- | --- | --- | --- | --- |
|  |  | **No. of Events/  Total No.** | **Incidence/ 1000 person-year** | **Absolute risk (%)** | **Crude** | | **Model 1** | | **Model 2** | | **No. of Events/  Total No.** | **Incidence/ 1000 person-year** | **Absolute risk (%)** | **Crude** | | **Model 2** | | **Model 2** | |
|  |  |  |  |  | **HR (95% CI)** | **P value** | **HR (95% CI)** | **P value** | **HR (95% CI)** | **P value** |  |  |  | **HR (95% CI)** | **P value** | **HR (95% CI)** | **P value** | **HR (95% CI)** | **P value** |
| **CAD PRS** | Low risk | 70/28502 | 0.27 (0.21 to 0.35) | 0.25 | Ref |  | Ref |  | Ref |  | 446/47087 | 1.07 (0.97 to 1.17) | 0.95 | Ref |  | Ref |  | Ref |  |
|  | Intermediate risk | 220/85868 | 0.29 (0.25 to 0.33) | 0.26 | 1.17 (0.86 to 1.58) | 0.32 | 1.06 (0.81 to 1.39) | 0.67 | 0.89 (0.59 to 1.35) | 0.58 | 1686/140899 | 1.35 (1.28 to 1.41) | 1.20 | 1.24 (1.12 to 1.37) | <0.001 | 1.26 (1.13 to 1.40) | 0.000 | 1.19 (1.02 to 1.40) | 0.03 |
|  | High risk | 110/27425 | 0.45 (0.37 to 0.54) | 0.40 | 1.81 (1.30 to 2.54) | 0.0004 | 1.64 (1.22 to 2.22) | 0.001 | 1.50 (0.95 to 2.38) | 0.08 | 611/44351 | 1.55 (1.43 to 1.68) | 1.38 | 1.44 (1.28 to 1.62) | <0.001 | 1.47 (1.30 to 1.66) | 0.000 | 1.34 (1.11 to 1.62) | 0.002 |
|  | Very high risk | 10/1451 | 0.77 (0.37 to 1.41) | 0.69 | 2.96 (1.41 to 6.21) | 0.004 | 2.85 (1.47 to 5.53) | 0.002 | 2.15 (0.64 to 7.23) | 0.22 | 57/2326 | 2.78 (2.10 to 3.60) | 2.45 | 2.57 (1.96 to 3.37) | <0.001 | 2.63 (2.00 to 3.47) | 0.000 | 1.90 (1.14 to 3.19) | 0.01 |
| **T2DM PRS** | Low risk | 58/28336 | 0.23 (0.17 to 0.30) | 0.20 | Ref |  | Ref |  | Ref |  | 496/47271 | 1.18 (1.08 to 1.29) | 1.05 | Ref |  | Ref |  | Ref |  |
|  | Intermediate risk | 258/86208 | 0.33 (0.29 to 0.38) | 0.30 | 1.66 (1.21 to 2.29) | 0.002 | 1.52 (1.14 to 2.01) | 0.004 | 1.57 (0.98 to 2.52) | 0.06 | 1658/140575 | 1.33 (1.27 to 1.39) | 1.18 | 1.11 (1.00 to 1.22) | 0.05 | 1.12 (1.01 to 1.23) | 0.02 | 1.11 (0.95 to 1.29) | 0.21 |
|  | High risk | 87/27257 | 0.36 (0.29 to 0.44) | 0.32 | 1.67 (1.15 to 2.42) | 0.007 | 1.62 (1.16 to 2.26) | 0.005 | 1.46 (0.84 to 2.52) | 0.18 | 596/44487 | 1.51 (1.39 to 1.64) | 1.34 | 1.28 (1.13 to 1.43) | <0.001 | 1.30 (1.15 to 1.46) | 0.000 | 1.23 (1.02 to 1.48) | 0.03 |
|  | Very high risk | 7/1445 | 0.54 (0.22 to 1.12) | 0.48 | 1.79 (0.64 to 4.96) | 0.27 | 2.46 (1.12 to 5.40) | 0.02 | 0.57 (0.08 to 4.33) | 0.59 | 50/2330 | 2.43 (1.80 to 3.21) | 2.15 | 2.10 (1.58 to 2.78) | <0.001 | 2.10 (1.57 to 2.81) | 0.000 | 1.96 (1.28 to 3.00) | 0.002 |

Model 1: Age + sex + genotyping array + first ten PC

Model 2: Age + sex + genotyping array + first ten PC + systolic BP + diastolic BP + BMI + waist circumference + current smoking + alcohol frequency + physical activity + eating habit

+ HbA1c + total cholesterol + triglyceride + HDL cholesterol + LDL cholesterol + lipoprotein (a) + estimated GFR + AST + ALT + Total bilirubin + albumin + white blood cell + hemoglobin + platelet + C-reactive protein

+ PRS specific disease + hypertension + dyslipidemia + heart failure + cancer + chronic kidney disease + stroke + chronic lung disease + chronic liver disease + Charlson comorbidity index

+ aspirin + lipid-lowering agent + anti-hypertensive agent

T2DM, type 2 diabetes mellitus; PRS polygenic risk score; PC, principal component; BP, blood pressure; GFR, glomerular filtration rate; AST, Aspartate aminotransferase; ALT, Alanine aminotransferase

**Supplementary Table 9. Hazard ratios and 95% confidential intervals for the cardiovascular mortality according to genetic risk and sex category**

| **CV mortality** | | **Male** | | | | | | | | | **Female** | | | | | | | | |
| --- | --- | --- | --- | --- | --- | --- | --- | --- | --- | --- | --- | --- | --- | --- | --- | --- | --- | --- | --- |
|  |  | **No. of Events/  Total No.** | **Incidence/ 1000 person-year** | **Absolute risk (%)** | **Crude** | | **Model 1** | | **Model 2** | | **No. of Events/  Total No.** | **Incidence/ 1000 person-year** | **Absolute risk (%)** | **Crude** | | **Model 1** | | **Model 2** | |
|  |  |  |  |  | **HR (95% CI)** | **P value** | **HR (95% CI)** | **P value** | **HR (95% CI)** | **P value** |  |  |  | **HR (95% CI)** | **P value** | **HR (95% CI)** | **P value** | **HR (95% CI)** | **P value** |
| **CAD PRS** | Low risk | 362/35020 | 1.16 (1.05 to 1.29) | 1.03 | Ref |  | Ref |  | Ref |  | 154/40569 | 0.43 (0.36 to 0.50) | 0.38 | Ref |  | Ref |  | Ref |  |
|  | Intermediate risk | 1400/104899 | 1.50 (1.42 to 1.58) | 1.33 | 1.29 (1.15 to 1.45) | <0.001 | 1.29 (1.15 to 1.45) | <0.001 | 1.22 (1.03 to 1.45) | 0.02 | 505/121809 | 0.46 (0.42 to 0.51) | 0.41 | 1.10 (0.91 to 1.31) | 0.32 | 1.10 (0.92 to 1.32) | 0.31 | 1.01 (0.74 to 1.37) | 0.95 |
|  | High risk | 536/33111 | 1.82 (1.67 to 1.98) | 1.62 | 1.57 (1.37 to 1.79) | <0.001 | 1.59 (1.39 to 1.81) | <0.001 | 1.47 (1.20 to 1.79) | <0.001 | 185/38665 | 0.54 (0.46 to 0.61) | 0.48 | 1.26 (1.02 to 1.56) | 0.03 | 1.27 (1.02 to 1.57) | 0.03 | 1.13 (0.78 to 1.63) | 0.53 |
|  | Very high risk | 50/1740 | 3.24 (2.41 to 4.28) | 2.87 | 2.78 (2.07 to 3.73) | <0.001 | 2.85 (2.12 to 3.83) | <0.001 | 2.33 (1.42 to 3.82) | <0.001 | 17/2037 | 0.94 (0.55 to 1.50) | 0.83 | 2.22 (1.34 to 3.66) | 0.002 | 2.27 (1.37 to 3.74) | 0.001 | 0.50 (0.07 to 3.66) | 0.50 |
| **T2D PRS** | Low risk | 397/34867 | 1.28 (1.16 to 1.41) | 1.14 | Ref |  | Ref |  | Ref |  | 157/40740 | 0.43 (0.37 to 0.50) | 0.39 | Ref |  | Ref |  | Ref |  |
|  | Intermediate risk | 1407/104888 | 1.51 (1.43 to 1.59) | 1.34 | 1.18 (1.06 to 1.32) | 0.004 | 1.20 (1.08 to 1.35) | 0.001 | 1.21 (1.02 to 1.43) | 0.03 | 507/121829 | 0.47 (0.43 to 0.51) | 0.42 | 1.08 (0.90 to 1.29) | 0.43 | 1.09 (0.91 to 1.30) | 0.36 | 1.03 (0.75 to 1.42) | 0.83 |
|  | High risk | 500/33238 | 1.70 (1.55 to 1.85) | 1.50 | 1.33 (1.17 to 1.52) | <0.001 | 1.37 (1.20 to 1.56) | <0.001 | 1.25 (1.02 to 1.53) | 0.03 | 183/38506 | 0.53 (0.46 to 0.62) | 0.48 | 1.24 (0.99 to 1.53) | 0.05 | 1.25 (1.01 to 1.55) | 0.04 | 1.33 (0.92 to 1.91) | 0.13 |
|  | Very high risk | 43/1770 | 2.75 (1.99 to 3.70) | 2.43 | 2.13 (1.55 to 2.92) | <0.001 | 2.22 (1.61 to 3.05) | <0.001 | 1.92 (1.22 to 3.04) | 0.005 | 14/2005 | 0.78 (0.43 to 1.32) | 0.70 | 1.79 (1.04 to 3.10) | 0.04 | 1.90 (1.10 to 3.28) | 0.02 | 1.54 (0.55 to 4.29) | 0.41 |

Model 1: Age + sex + genotyping array + first ten PC

Model 2: Age + sex + genotyping array + first ten PC + systolic BP + diastolic BP + BMI + waist circumference + current smoking + alcohol frequency + physical activity + eating habit

+ HbA1c + total cholesterol + triglyceride + HDL cholesterol + LDL cholesterol + lipoprotein (a) + estimated GFR + AST + ALT + Total bilirubin + albumin + white blood cell + hemoglobin + platelet + C-reactive protein

+ PRS specific disease + hypertension + dyslipidemia + heart failure + cancer + chronic kidney disease + stroke + chronic lung disease + chronic liver disease + Charlson comorbidity index

+ aspirin + lipid-lowering agent + anti-hypertensive agent

T2DM, type 2 diabetes mellitus; PRS polygenic risk score; PC, principal component; BP, blood pressure; GFR, glomerular filtration rate; AST, Aspartate aminotransferase; ALT, Alanine aminotransferase

**Supplementary Table 10. Hazard ratios and 95% confidential intervals for the cardiovascular mortality according to genetic risk and prevalent disease of type 2 diabetes mellitus or coronary artery disease at baseline**

| **CV mortality** | | **T2DM at baseline** | | | | | | | | | **CAD at baseline** | | | | | | | | |
| --- | --- | --- | --- | --- | --- | --- | --- | --- | --- | --- | --- | --- | --- | --- | --- | --- | --- | --- | --- |
|  |  | **No. of Events/  Total No.** | **Incidence/ 1000 person-year** | **Absolute mortality rate** | **Crude** | | **Model 1** | | **Model 2** | | **No. of Events/  Total No.** | **Incidence/ 1000 person-year** | **Absolute mortality rate** | **Crude** | | **Model 1** | | **Model 2** | |
|  |  |  |  |  | **HR (95% CI)** | **p value** | **HR (95% CI)** | **p value** | **HR (95% CI)** | **p value** |  |  |  | **HR (95% CI)** | **p value** | **HR (95% CI)** | **p value** | **HR (95% CI)** | **p value** |
| **CAD PRS** | Low risk | 70/2725 | 2.91 (2.26 to 3.67) | 2.57 | Ref |  | Ref |  | Ref |  | 120/2950 | 4.62 (3.83 to 5.53) | 4.07 | Ref |  | Ref |  | Ref |  |
|  | Intermediate risk | 266/9030 | 3.34 (2.95 to 3.77) | 2.95 | 1.22 (0.94 to 1.57) | 0.14 | 1.23 (0.95 to 1.59) | 0.12 | 0.88 (0.59 to 1.32) | 0.55 | 547/12592 | 4.95 (4.54 to 5.39) | 4.34 | 1.14 (0.96 to 1.35) | 0.14 | 1.11 (0.94 to 1.32) | 0.23 | 1.19 (0.90 to 1.58) | 0.22 |
|  | High risk | 120/3099 | 4.40 (3.65 to 5.26) | 3.87 | 1.65 (1.23 to 2.21) | 0.0008 | 1.71 (1.28 to 2.29) | 0.0003 | 1.14 (0.71 to 1.84) | 0.58 | 247/5639 | 5.00 (4.40 to 5.67) | 4.38 | 1.16 (0.94 to 1.43) | 0.15 | 1.15 (0.93 to 1.42) | 0.19 | 1.43 (1.02 to 2.01) | 0.04 |
|  | Very high risk | 14/180 | 8.94 (4.89 to 15.00) | 7.78 | 3.60 (1.99 to 6.48) | <0.001 | 3.86 (2.14 to 6.98) | <0.001 | 4.16 (1.56 to 11.10) | 0.004 | 24/497 | 5.51 (3.53 to 8.20) | 4.83 | 1.19 (0.63 to 2.24) | 0.60 | 1.19 (0.63 to 2.25) | 0.59 | 0.66 (0.09 to 4.83) | 0.69 |
| **T2DM PRS** | Low risk | 50/1758 | 3.22 (2.39 to 4.25) | 2.84 | Ref |  | Ref |  | Ref |  | 142/3675 | 4.39 (3.70 to 5.17) | 3.86 | Ref |  | Ref |  | Ref |  |
|  | Intermediate risk | 264/8724 | 3.43 (3.03 to 3.87) | 3.03 | 1.16 (0.91 to 1.49) | 0.23 | 1.20 (0.94 to 1.54) | 0.14 | 1.34 (0.88 to 2.04) | 0.18 | 559/13183 | 4.83 (4.44 to 5.25) | 4.24 | 1.19 (0.99 to 1.41) | 0.06 | 1.20 (1.01 to 1.43) | 0.04 | 1.25 (0.93 to 1.67) | 0.14 |
|  | High risk | 138/4222 | 3.71 (3.12 to 4.38) | 3.27 | 1.19 (0.88 to 1.60) | 0.26 | 1.23 (0.92 to 1.66) | 0.17 | 1.16 (0.69 to 1.95) | 0.58 | 211/4529 | 5.32 (4.63 to 6.10) | 4.66 | 1.35 (1.09 to 1.66) | 0.005 | 1.37 (1.11 to 1.69) | 0.003 | 1.37 (0.97 to 1.93) | 0.08 |
|  | Very high risk | 18/330 | 6.23 (3.69 to 9.85) | 5.45 | 2.67 (1.42 to 5.01) | 0.002 | 2.99 (1.59 to 5.62) | 0.0006 | 2.82 (1.03 to 7.72) | 0.04 | 26/291 | 10.42 (6.81 to 15.27) | 8.93 | 2.19 (1.33 to 3.61) | 0.002 | 2.31 (1.40 to 3.80) | 0.001 | 2.46 (1.11 to 5.46) | 0.03 |

Model 1: Age + sex + genotyping array + first ten PC

Model 2: Age + sex + genotyping array + first ten PC + systolic BP + diastolic BP + BMI + waist circumference + current smoking + alcohol frequency + physical activity + eating habit

+ HbA1c + total cholesterol + triglyceride + HDL cholesterol + LDL cholesterol + lipoprotein (a) + estimated GFR + AST + ALT + Total bilirubin + albumin + white blood cell + hemoglobin + platelet + C-reactive protein

+ PRS specific disease + hypertension + dyslipidemia + heart failure + cancer + chronic kidney disease + stroke + chronic lung disease + chronic liver disease + Charlson comorbidity index

+ aspirin + lipid-lowering agent + anti-hypertensive agent

T2DM, type 2 diabetes mellitus; PRS polygenic risk score; PC, principal component; BP, blood pressure; GFR, glomerular filtration rate; AST, Aspartate aminotransferase; ALT, Alanine aminotransferase

**Supplementary Figure 1. Density and prevalence plot according to the PRS distribution (A) CAD, (B) T2DM**

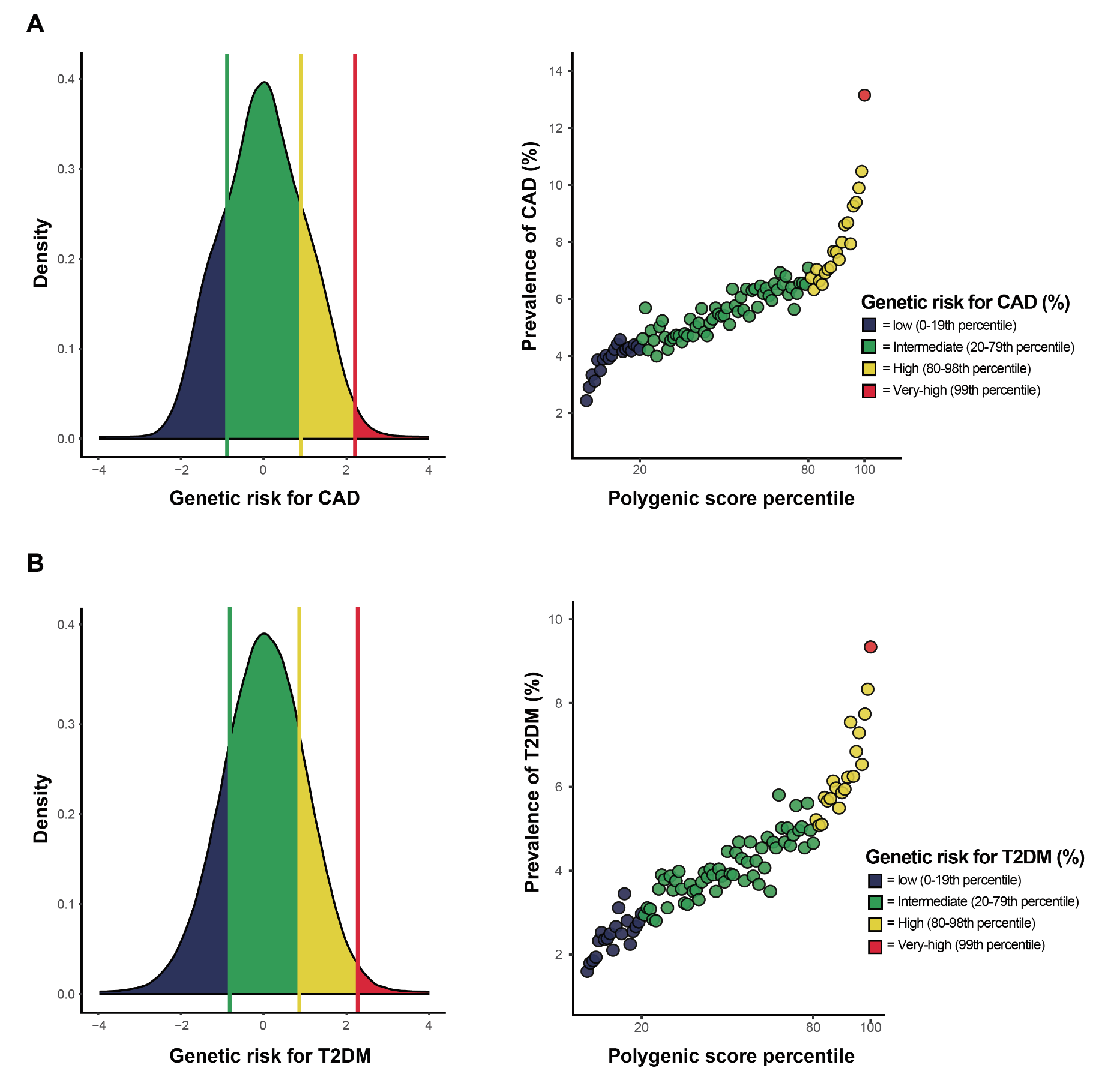

PRS, polygenic risk score; CAD, coronary artery disease; T2DM, type 2 diabetes mellitus

**Supplementary Figure 2. 10-year cardiovascular mortality rates according to (A) genetic risk for CAD, (B) genetic risk for T2DM and lifestyle behavior**

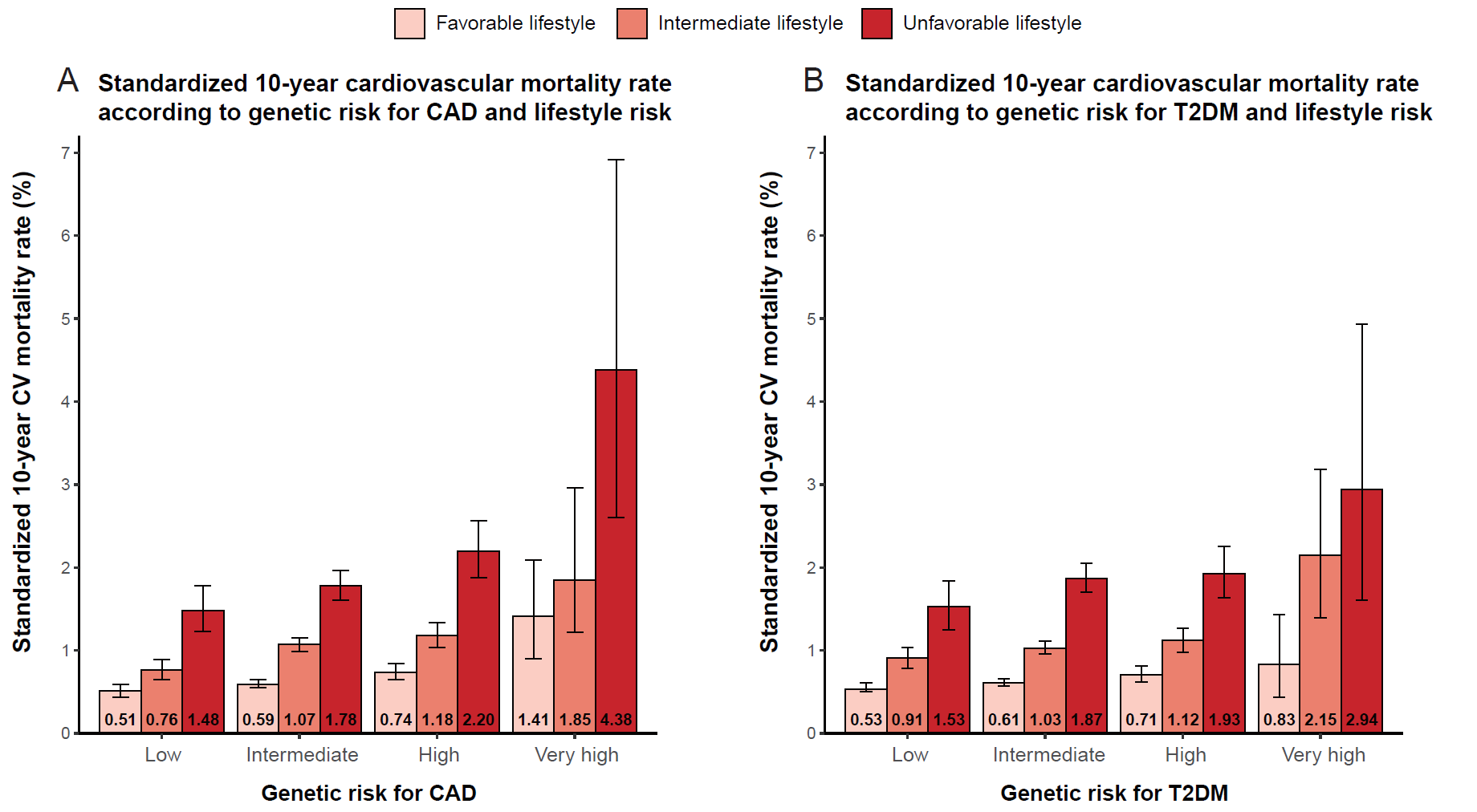

CV, cardiovascular; CAD, coronary artery disease; T2DM, type 2 diabetes mellitus

**Supplementary Figure 3. Forest plot of cardiovascular mortality according to genetic risk for CAD and four lifestyle behaviors**

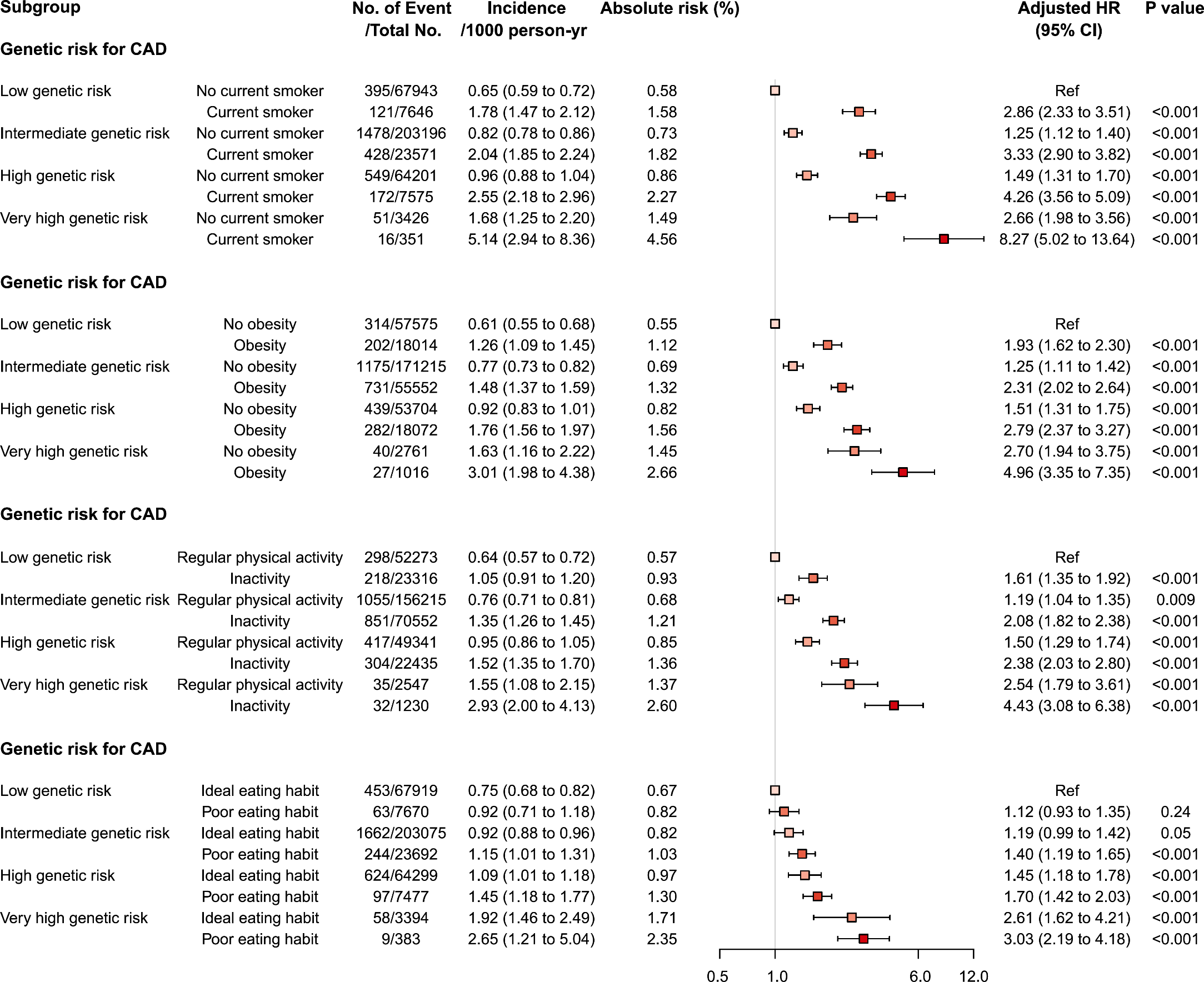

CAD, coronary artery disease

**Supplementary Figure 4. Forest plot of cardiovascular mortality according to genetic risk for T2DM and four lifestyle behaviors**

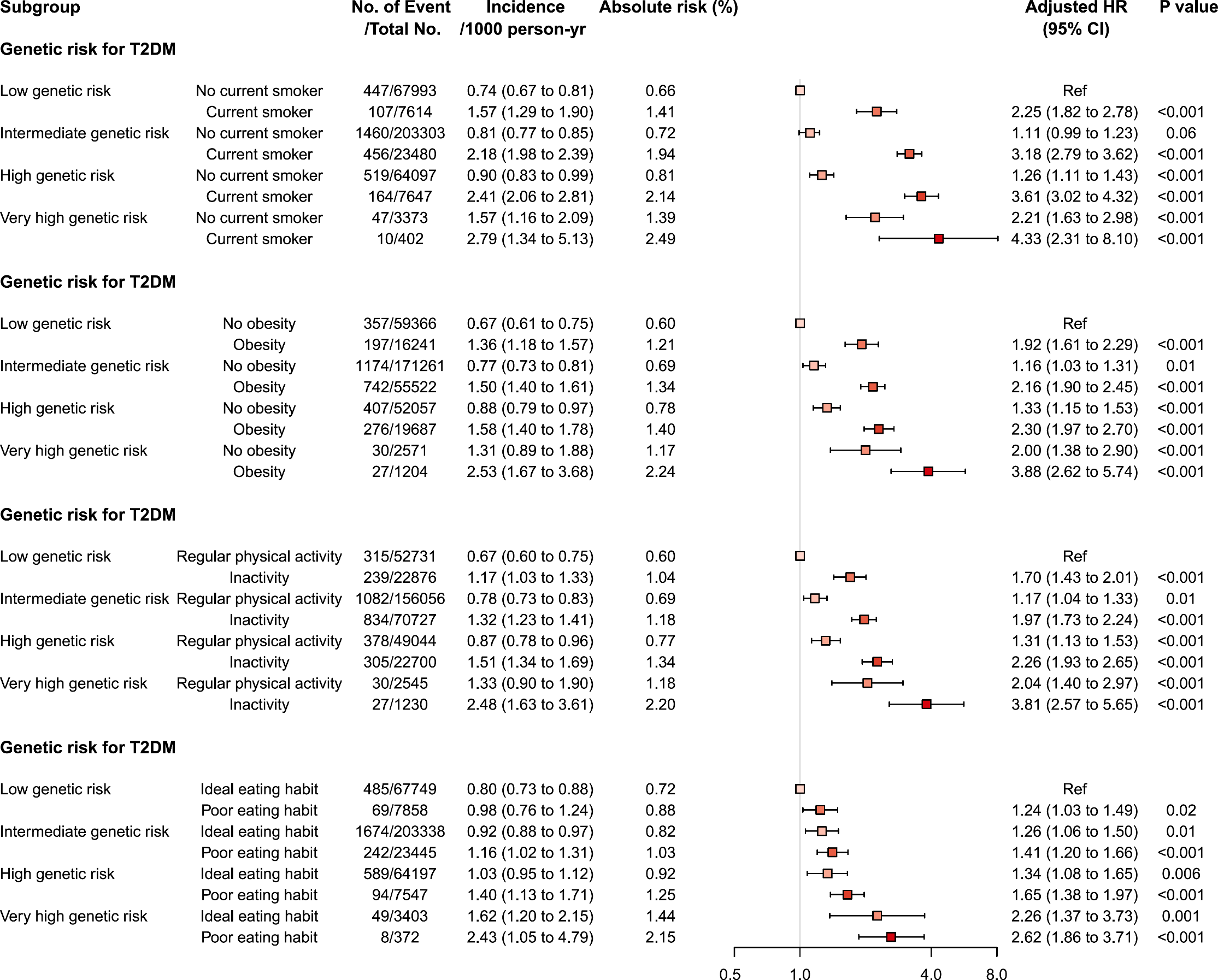

T2DM, type 2 diabetes mellitus

**Supplementary Figure 5. 10-year cardiovascular mortality rates according to (A) genetic risk for CAD and age categories, (B) genetic risk for T2DM and age categories, (C) genetic risk for CAD and sex categories, (D) genetic risk for T2DM and sex categories**

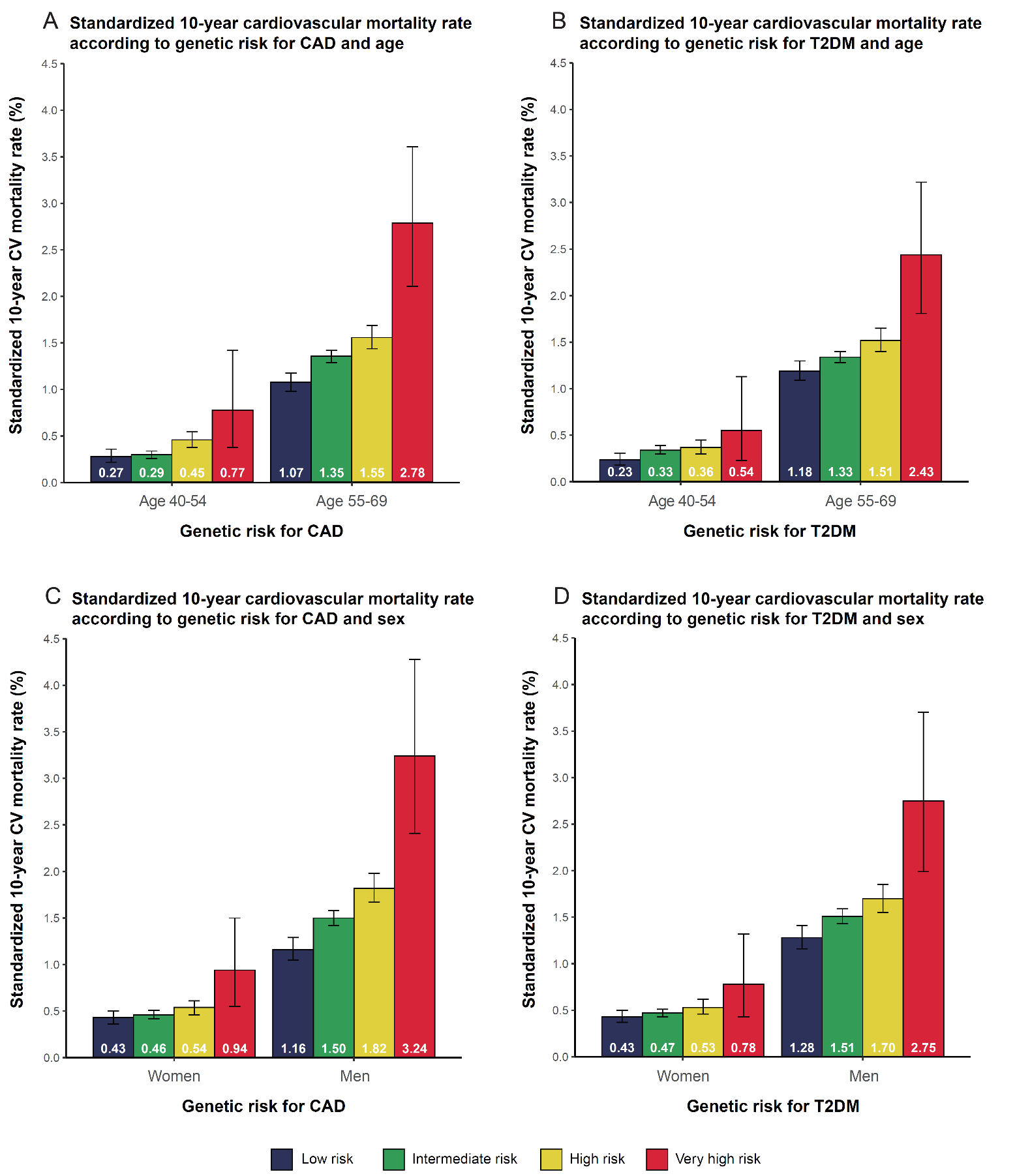

CV, cardiovascular; CAD, coronary artery disease; T2DM, type 2 diabetes mellitus

**Supplementary Figure 6. 10-year cardiovascular mortality rates according to genetic risk and prevalent disease status; (A) genetic risk for CAD and CAD history at baseline, (B) genetic risk for CAD and T2DM history at baseline, (C) genetic risk for T2DM and CAD history at baseline, (D) genetic risk for T2DM and T2DM history at baseline**

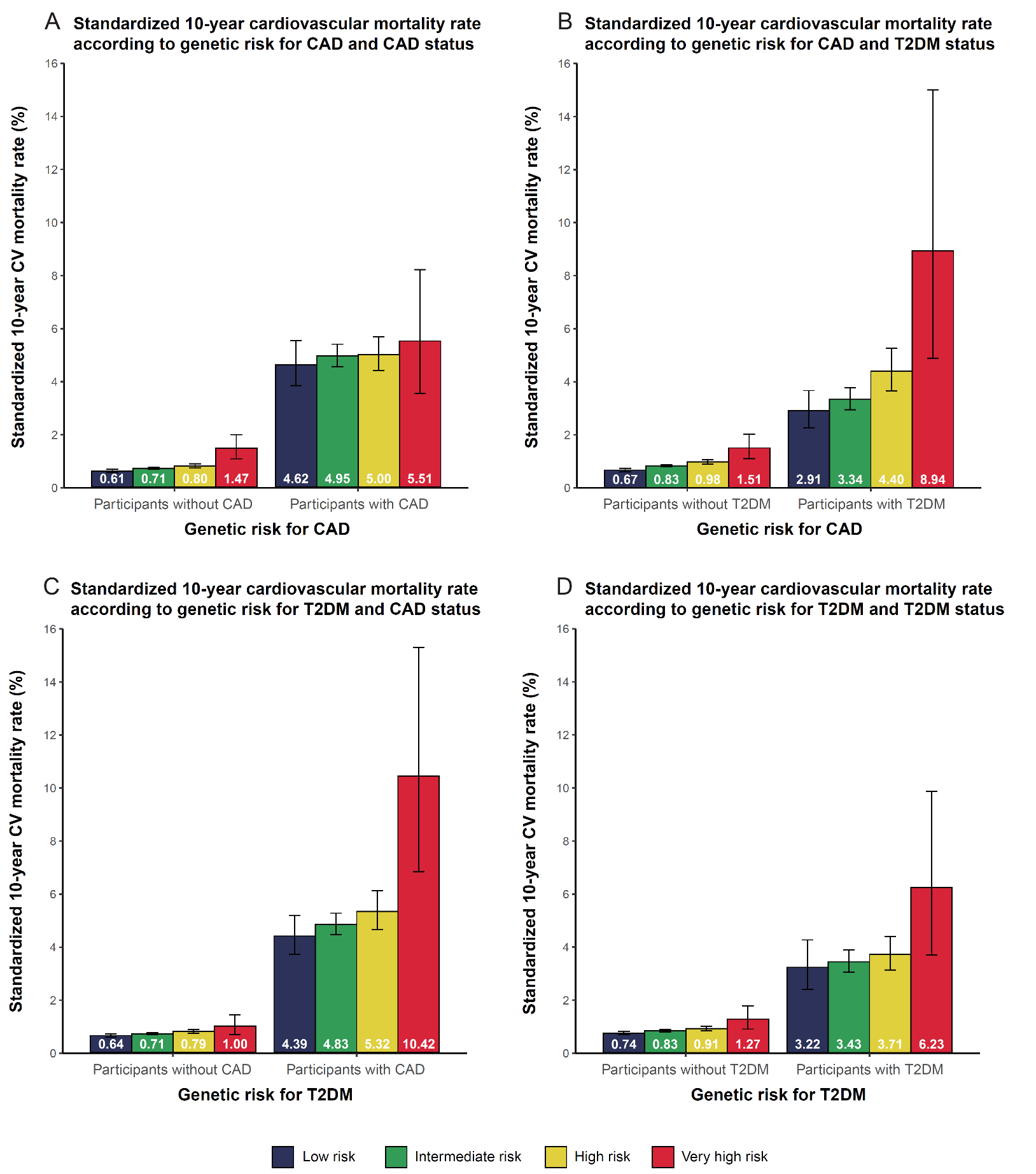

CV, cardiovascular; CAD, coronary artery disease; T2DM, type 2 diabetes mellitus

**Supplementary Figure 7. Forest plot of cardiovascular mortality according to genetic risk and sex in patients with diabetes at baseline**

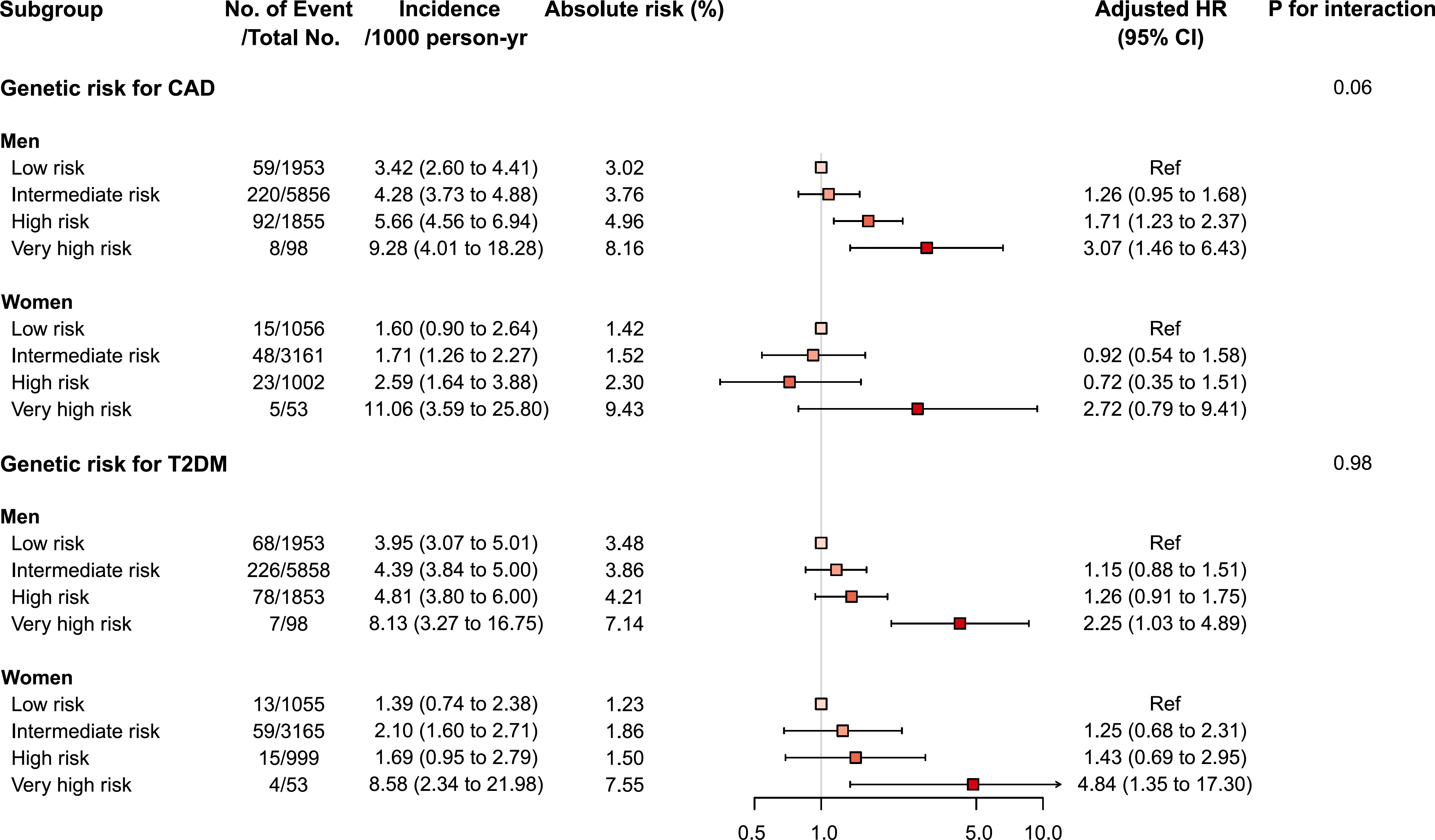

CAD, coronary artery disease; T2DM, type 2 diabetes mellitus

P values are for testing the interaction between each genetic risk category and sex.
